# Evaluation of the effectiveness of remdesivir in severe COVID-19 using observational data from a prospective national cohort study

**DOI:** 10.1101/2021.06.18.21259072

**Authors:** B N Arch, D Kovacs, J T Scott, A P Jones, E M Harrison, A Rosala-Hallas, C G Gamble, P J M Openshaw, J K Baillie, M G Semple, ISARIC4C Investigators

## Abstract

**Background:** Remdesivir has been evaluated in clinical trial populations, but there is a sparsity of evidence evaluating effectiveness in general populations.

**Methods:** Adults eligible to be treated with remdesivir, requiring oxygen but not ventilated, were identified from UK patients hospitalised with COVID-19. Patients treated with remdesivir within 24h of hospitalisation were compared with propensity-score matched controls; estimates of effectiveness were calculated for short-term outcomes (14-day mortality, 28-day mortality, time-to-recovery among others) using multivariable modelling.

**Results:** 9,278 out of 39,330 patients satisfied eligibility criteria. 1,549 patients were identified as ‘treated’ and matched with 4,964 controls. Patients were 62% male, mean (SD) age 63.1 (15.6) years, 80% ‘White’ ethnicity, and symptomatic for a median of 6 days prior to baseline. There was no statistically significant benefit of remdesivir at 14 days in terms of mortality or clinical status; there were signals of effectiveness in time-to-recovery after day 9, and a reduction in 28-day mortality.

**Conclusion:** In a real-world setting, initiation of remdesivir within 24h of hospitalisation in conjunction with standard of care was not associated with a benefit at 14 days but supports clinical trial evidence of a potential reduction in 28-day mortality.

## Background

Several therapeutic drugs licensed for use in other conditions have been trialled in the treatment of severe COVID-19. Remdesivir (GS-5734), was given emergency approval for use on 26^th^ May 2020 in people aged 12 years and older affected with severe COVID-19 by the United Kingdom’s (UK) Medicines and Healthcare products Regulatory Agency (MHRA) and commissioned for routine use in severe COVID-19 following an evidence review [1] by the National Institute for Health and Care Excellence (NICE) in July 2020. Evaluation of its efficacy in this UK population was sought.

### Clinical trial evidence for remdesivir use

Remdesivir is a broad-spectrum antiviral drug that has shown activity against Ebola virus *in vitro* and in non-human primates [2]. It is an adenosine nucleotide prodrug administered via intravenous infusion, and once it is metabolised into its active form [3], it inhibits the viral RNA-dependent RNA polymerase [4], a conserved enzyme involved in viral RNA synthesis. Remdesivir has demonstrated *in vitro* efficacy against other emerging coronaviruses, such as MERS-CoV and SARS-CoV-1 [5, 6], and SARS-CoV-2 [7-11]. The half-effective concentration (EC_50_) values against SARS-CoV-2 were below 5µM. This promising EC_50_ combined with high (>100) safety indices in cells, made remdesivir one of the principal compounds of interest for a clinical trial early in the pandemic. While *in vivo* studies also showed clinical benefits [2, 12], there are limits to what can be extrapolated from the animal models due to important differences in the pharmacokinetics of the drug and disease course, particularly in mice [13]. The pharmacokinetics of remdesivir have been reported in healthy adults, showing a favourable profile [14], but they are yet to be reported in severely ill patients. Four clinical trials have published results: two small trials [15, 16], ACTT-1 [17], and SOLIDARITY [18]. A meta-analysis of 28-day mortality results from these is presented in the SOLIDARITY paper [18]: an overall rate-ratio for death of 0.91 (95% CI, 0.79 to 1.05), indicating no significant reduction in mortality because of remdesivir use. Of note, there was a signal of greater potential for benefit in non-ventilated patients, 0.80 (0.63–1.01). A striking benefit was reported for 14-day mortality in ACTT-1, though this was a secondary outcome of that trial.

## Methods

### Study design and participants

Our study used data from a prospective cohort of UK patients hospitalised with COVID-19: the International Severe Acute Respiratory and emerging Infections Consortium (ISARIC) World Health Organization (WHO) Clinical Characterisation Protocol UK (CCP-UK), implemented by the ISARIC Coronavirus Clinical Characterisation Consortium (ISARIC-4C) in 260 hospitals across England, Scotland, and Wales. The protocol and further study details are available online [19]. ‘Baseline’ was defined as ‘date of hospital admission’ for community acquired COVID-19 and ‘date of a positive COVID-19 test’ for hospital acquired infection. Patients were followed up for 28 days post baseline. The study period was patient baseline between 26^th^ May 2020 and 30^th^ November 2020. Under the Control of Patient Information (COPI) notice 2020 for urgent public health research, processing of demographic and routine clinical data from medical records for research does not require consent in England and Wales. In Scotland, a waiver for consent was obtained from the Public Benefit and Privacy Panel.

Inclusion and exclusion criteria were used to identify a subgroup of patients that would have been eligible to have received remdesivir, and not initially ventilated during the first 24h post baseline. Propensity-score methods were applied to identify a control group for whom the likelihood of being given remdesivir was of a similar distribution to the treatment group, and to balance for baseline factors that are related to underlying risk of 14-day mortality. We assessed effectiveness of remdesivir by comparing these groups with respect to several outcomes. We have followed the Strengthening the Reporting of Observational Studies in Epidemiology (STROBE) Statement checklist to guide transparent reporting of this study.

Patients eligible to have received remdesivir were identified as satisfying four inclusion criteria: (1) Laboratory confirmed SARS-CoV-2 infection; (2) hospitalised; (3) aged ≥ 18 years at baseline; (4) requiring supplementary oxygen (usually for hypoxaemia SpO_2_ ≤ 94%) at any time during the 24h post baseline; and not confirmed pregnant or suffering from chronic kidney disease (CKD). We further excluded patients with (1) evidence of requiring any ventilation within 24h of baseline (high flow oxygen, non-invasive, invasive ventilation, or extracorporeal membrane oxygenation (ECMO); (2) previous hospitalisation due to COVID-19; (3) no baseline CRF; (4) a missing ‘Medication’ section of the Treatment CRF; (4) remdesivir initiated > 24h post baseline.

We define two treatment groups: standard of care with remdesivir initiated within 24hrs of baseline (remdesivir group); and standard of care without remdesivir (control group). Guidelines in the UK [20] – recommended remdesivir’s use in newly hospitalised patients, no later than within 10 days of symptom onset, and only initiated in non-ventilated patients. Recommended dosing was 200mg on Day 1 of treatment, followed by 100mg daily for 4 days. Guidelines regarding recommended duration of treatment (see Supplementary Material) and standard of care evolved over time: corticosteroids, dexamethasone, and hydrocortisone, became recommended for some patients part-way through the study period. Interactions between remdesivir and these corticosteroids were not expected such that remdesivir patients will have received these in the same way as non-remdesivir patients. The drugs hydroxychloroquine and chloroquine phosphate were actively not recommended as concomitant medications for remdesivir from 3^rd^ September 2020 (Figure 1).

**Figure 1:**
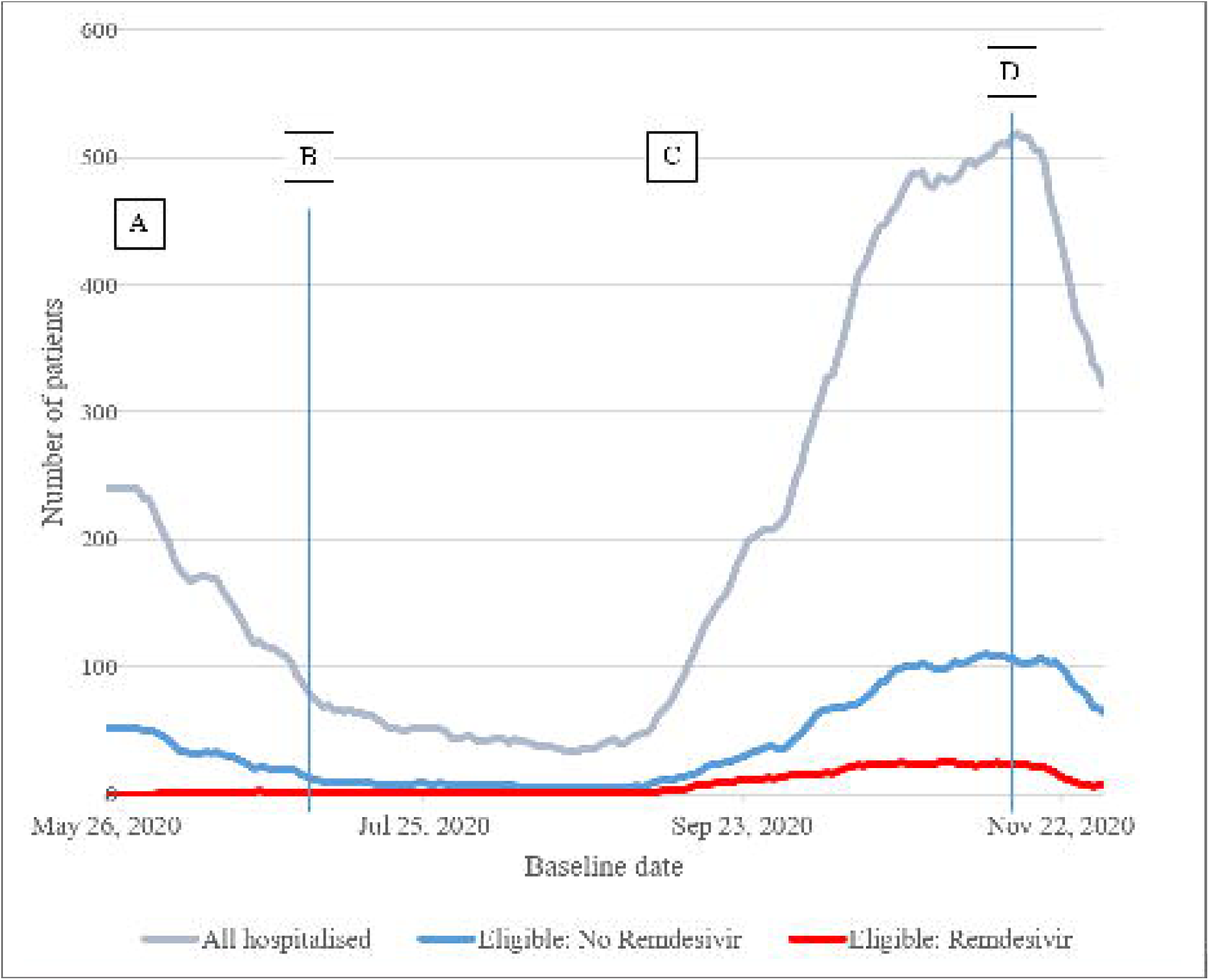
Number of patients hospitalised in the UK due to COVID-19, recorded in ISARIC4C Database, and of these the number eligible for inclusion in this study (7-day rolling average) **Footnote for Figure 1** <u>Key time-points</u>: A: Early access to medicine (EAMS) starts; B: EAMS ends and remdesivir commissioned for routine use in the UK; C: guidelines given for co-administration of corticosteroids; D: guidelines implemented ‘consider stopping remdesivir if: the patient clinically improves and no longer requires supplemental oxygen 72 h after commencement of treatment; or the patient continues to deteriorate despite 48 h of sustained mechanical ventilation.’

Data were routinely collected on patients at baseline, first day of admission to an ICU, and on death, discharge, or day 28 depending on which was soonest. For some sites, daily CRFs for days 3, 6 and 9 were also collected. Remdesivir patients had daily CRFs completed for each day of remdesivir dosing, and on day 14 after remdesivir initiation. Data collection regarding safety was limited to a tick-box assessment of 30 complications during hospital stay post baseline, and a free-text entry of other complications. Free text was used to identify other commonly occurring complications. There was no scope to measure severity or relatedness, and the quality of the data relied on what was recorded in medical notes. Missingness: CRFs could be partially completed, or entirely missing. A missing Day 1 and/or Treatment CRF represented an exclusion criterion; categorical data missing from partially completed CRFs were handled by using ‘Unknown’ as a category. Interpolation was not used to handle missingness in continuous data. An extract of the database was made prior to full analysis to assess (a) whether the sample size of eligible remdesivir patients was >500, and (b) the extent of missing data of key baseline variables and the primary outcome.

We specified a primary outcome in our statistical analysis plan to be 14-day mortality, as this time-point is important in ACTT-1 but not reported by SOLIDARITY. A number of other outcomes were also specified: (1) time-to-recovery (discharge from hospital or continued hospitalisation with no on-going health-care needs related to COVID-19); (2) 28-day mortality; (3) time-to-death; (4) clinical status at day 15; (5) length of time receiving supplementary oxygen; (6) time-to-first ventilation; (7) use of non-invasive ventilation; (8) use of mechanical ventilation or ECMO; (9) acute renal injury/acute renal failure; and (10) liver dysfunction. Full definitions and rationale for these outcomes are given in the Supplementary Material.

### Statistical Methods

The study was planned prior to access to the data being granted and a statistical analysis plan (SAP) was published on the ISARIC website on 16^th^ December [21]. This was subject to internal clinical and statistical review. The manufacturer was given opportunity to comment on the SAP - this was discretionary, and suggested revisions were considered prior to finalisation; none were considered substantive, and the proposed methodology was unchanged.

A formal sample size calculation was not undertaken. No upper boundary was placed on sample size. The number of controls eligible for the study could not be predicted, but the total was expected to be greater than the number that received remdesivir.

Likelihood of receiving remdesivir according to patients’ baseline characteristics was modelled using logistic regression. Baseline factors were prespecified in the SAP, chosen to be potentially associated with likelihood of receiving remdesivir, or known *a priori* to be associated with short-term mortality [22]. These were: month of baseline; ISARIC4C tier of participating centre (0/1/2); sex; age; broad ethnicity group (White/Asian/Black/Other); clinically extremely vulnerable status (Yes [Any of the following: cancer, severe respiratory condition, on immunosuppression therapy, other]/None/ Unknown); diabetes; hypertension; obesity; chronic cardiac disease (CCD); chronic pulmonary disease (CPD); asthma; where COVID-19 was acquired (community/hospital); admitted to HDU or ICU at baseline (Yes/No). Several models were fitted (see Supplementary Material) using different strategies of model selection. Propensity scores were calculated under each model, and a potential set of controls selected: each remdesivir patient was matched to up to four controls with a similar propensity score (calliper-width 0.2 standard deviations [23]); controls were chosen without replacement. Underlying probability of death at 14-days was modelled using logistic regression in eligible non-remdesivir patients and used to calculate a disease risk score for each patient in the study. A balance statistic was calculated for each propensity score model using weighted standardised difference in risk score (ASD_RS_). An optimal propensity score model was selected minimising ASD_RS_. A pre-defined balance diagnostic threshold: ASD_RS_ ≤ 0.1 [24] was used to indicate adequate balance. Further details and rationale for methods are given in supplementary material.

Descriptive statistics were used to describe demographics, other baseline characteristics including pre-existing comorbidities, treatments received during hospitalisation, and complications associated with hospitalisation. Outcomes were summarised by treatment group, but inference was obtained through multivariable modelling, adjusting for confounders to increase robustness. Confounders chosen *a priori* (sex, age-group, and number of key comorbidities) were identified by Knight *et al* [22] as being key underlying predictors of short-term mortality in COVID-19. We allowed for additional factors to be added to outcome models after seeing the data. Models incorporated weights for the control group to account for the variable matching ratio (*w*_*j*_ = 1/*k*_*j*_) where *k*_*j*_ is the number of controls matched to remdesivir patient *j*). Stuart [23] suggests that for analysis purposes, the two groups may be treated as independent. Binary outcomes were modelled using logistic regression; time-to-event outcomes with Cox Proportional-Hazards modelling (partitioning the time axis and estimating hazard-ratios within sub-intervals should proportional hazards not hold); ordinal outcomes with ordinal logistic regression. The primary outcome analysis was subject to sensitivity analyses regarding specification of the logistic regression model. Propensity score matching was carried out using *MatchIt* [25] in R v3.6.1; all other analyses were carried out using SAS v9.4.

## Results

### Cohort ascertainment and characteristics

A total of 39,330 unique patients were identified from a data extract made on 8^th^ January 2021, with a baseline date between 26^th^ May and 30^th^ November 2020. This extract was judged to be adequate for the purpose of our analysis: a sample size of approximately 1,500 remdesivir patients, over 7,700 potential controls, and <5% patients with missing primary outcome. A total of 10,434 patients satisfied inclusion/exclusion criteria. Of these, 1,156 had remdesivir initiated > 24h after baseline, and could not be included in the study, leaving an eligible cohort of 9,278.

An optimal propensity score model was derived, with sufficient balance between groups (ASD_RS_= 0.01). In the eligible cohort, remdesivir within 24h of baseline was found to have been more likely given: later in the study period (Sept-Nov); to younger patients; those without an extreme clinical vulnerability; with obesity; who acquired COVID-19 in the community; or who were not admitted to HDU or ICU at baseline (Supplementary Table 5). The fitted primary outcome risk score model results are presented in Supplementary Table 3. The cohorts are well balanced with respect to baseline characteristics (Supplementary Table 5, Supplementary Figure 2) – in particular, 4C Mortality Score [22] distributions were almost identical.

6,513 were included in the matched analysis (Figure 2). The median (IQR) number of controls per remdesivir patient was 3 (3, 4). Patients included in the matched cohort came from all regions of the UK, were 62.1% male, with mean (SD) age 63.1 (15.6) years, and 79.7% of those with ethnicity recorded were categorised as ‘White’ (Table 1). They had a median (IQR) 4C Mortality Score of 9 (6, 12), meaning that most patients were classified as at intermediate or high risk of in-hospital mortality.

**Figure 2:**
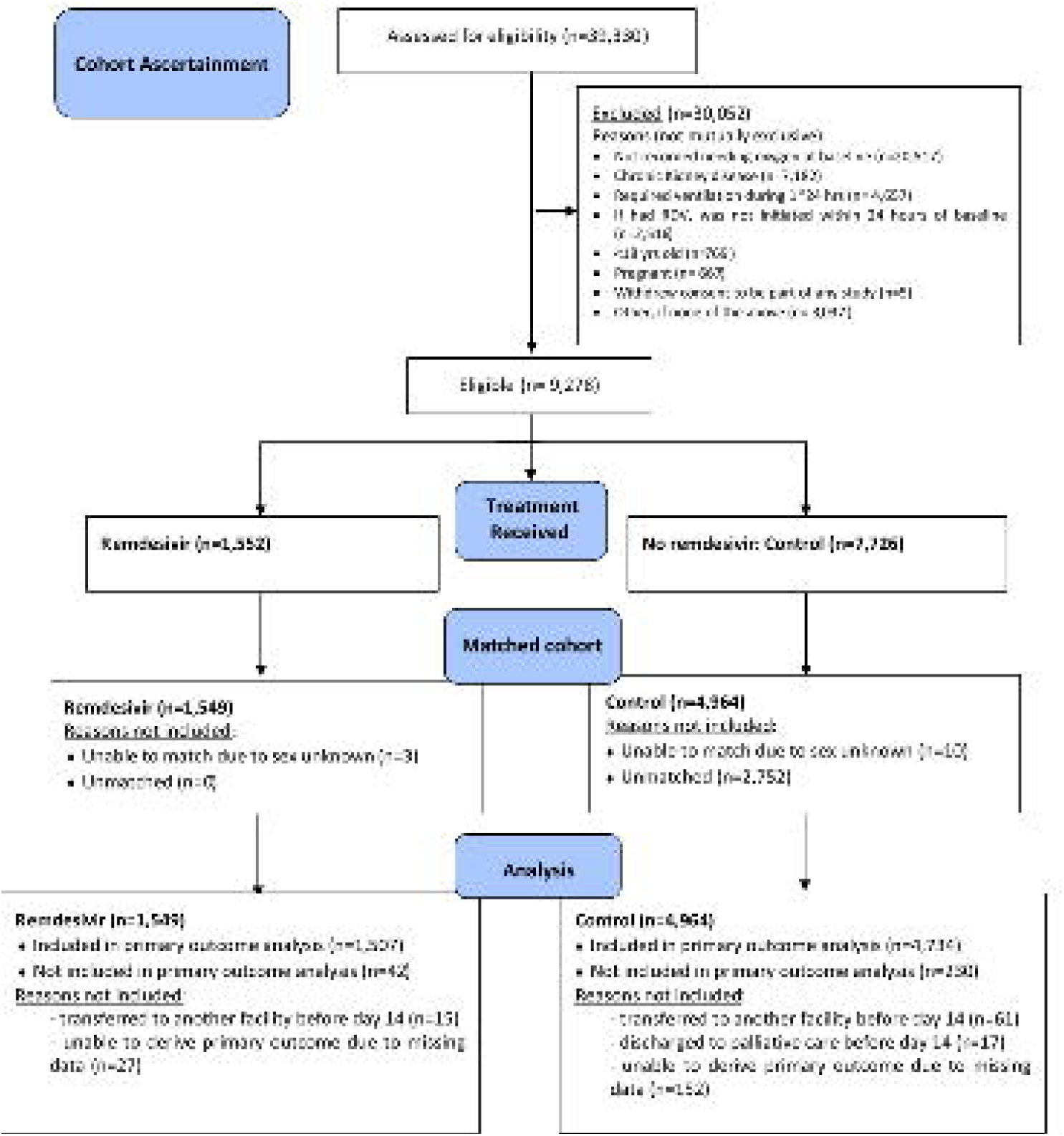
CONSORT style flow-chart summarising the flow of patients through the analysis stages

**Table 1.**
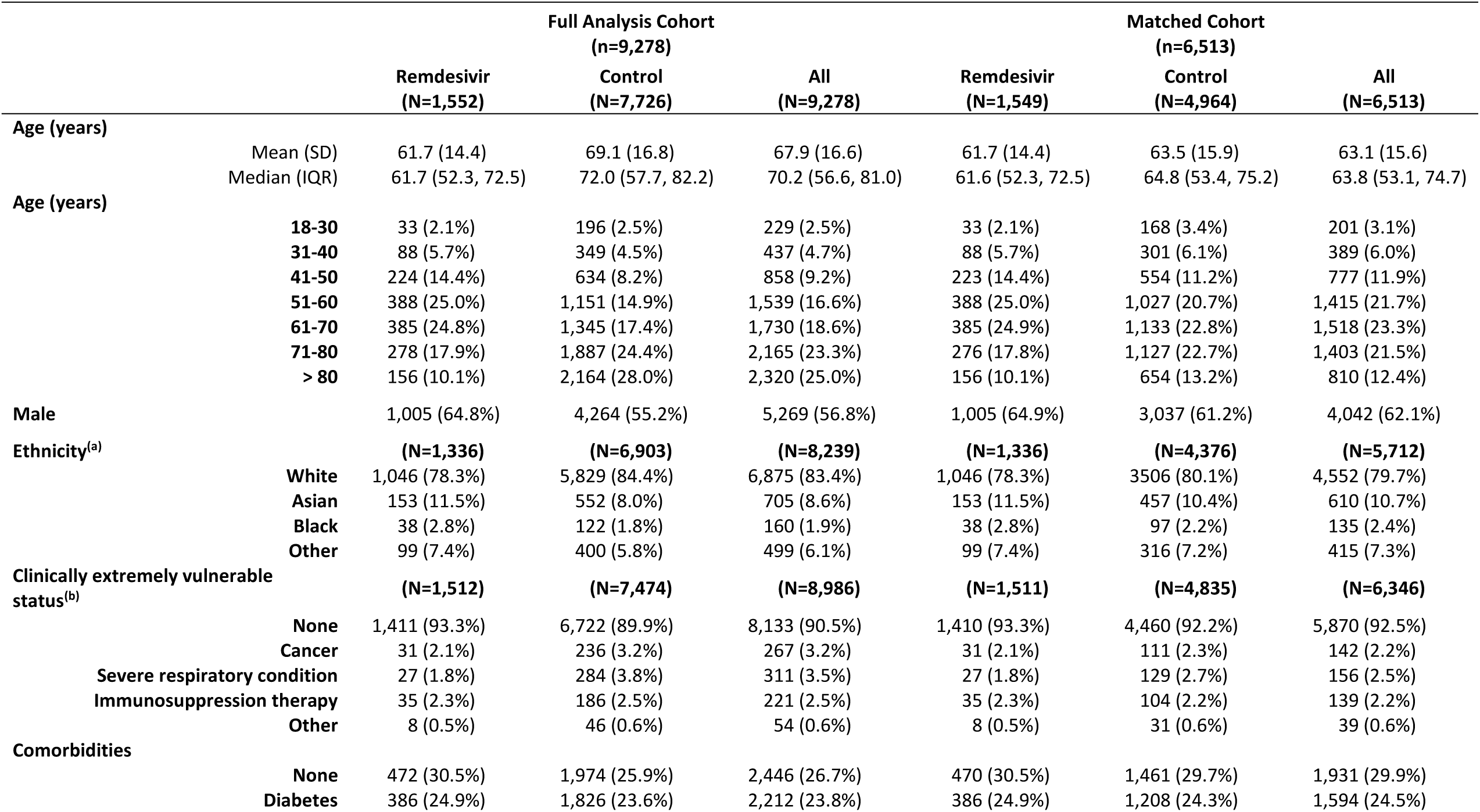
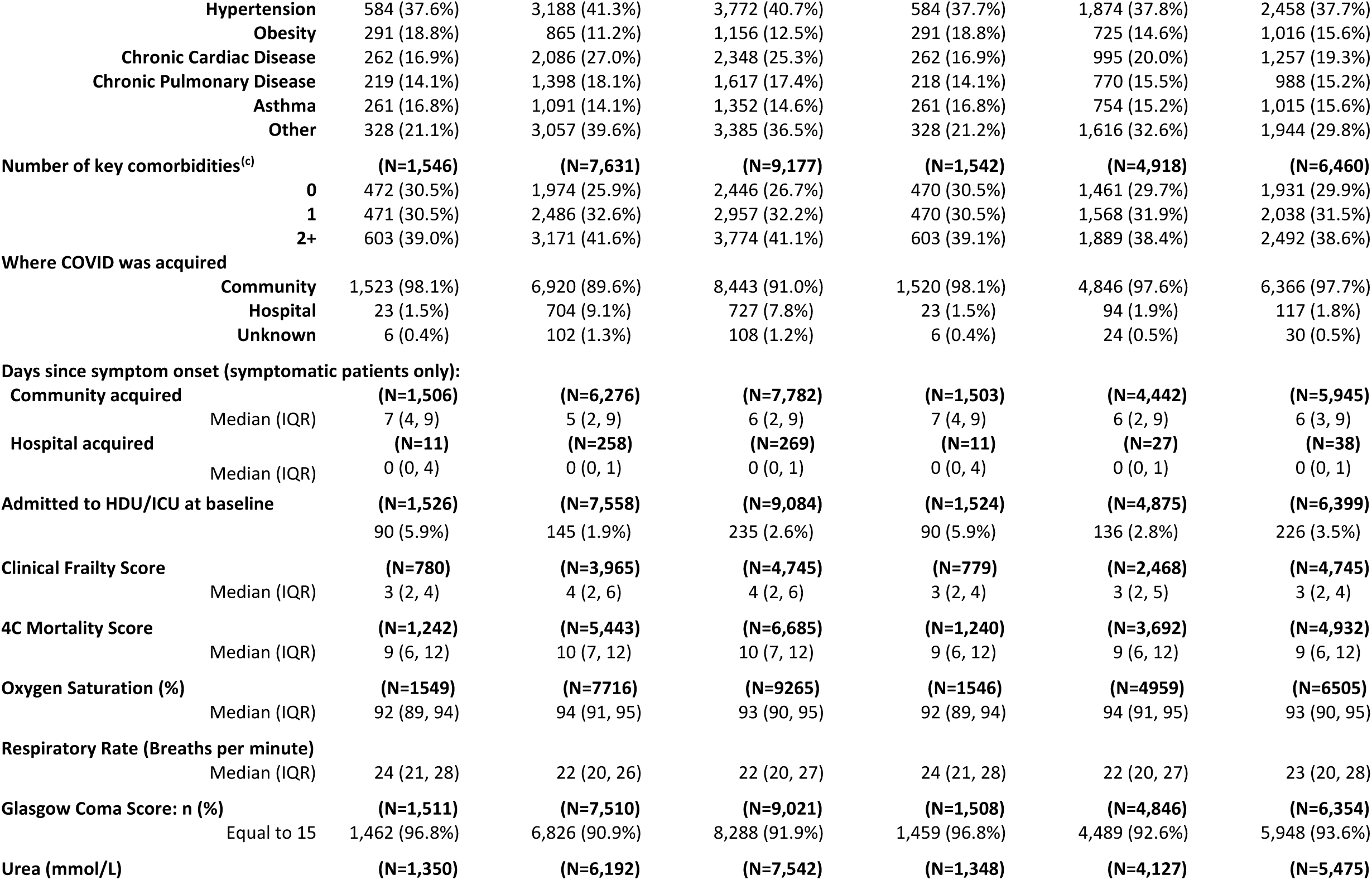
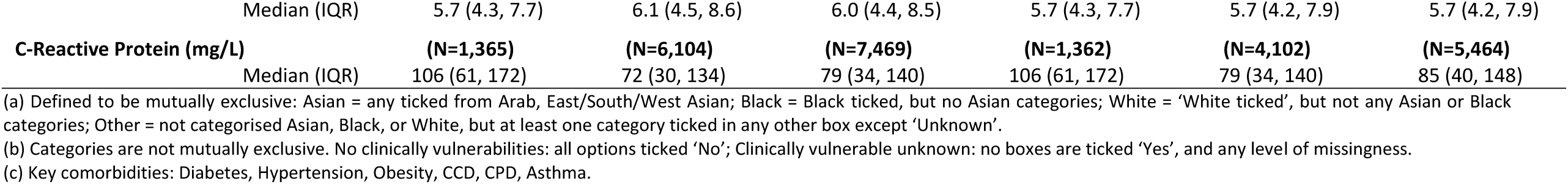
Baseline characteristics (statistics are n(%) unless otherwise stated)

|  |  | Full Analysis Cohort<br>(n=9,278) |  |  | Matched Cohort<br>(n=6,513) |  |  |
| --- | --- | --- | --- | --- | --- | --- | --- |
|  |  | Remdesivir<br>(N=1,552) | Control<br>(N=7,726) | All<br>(N=9,278) | Remdesivir<br>(N=1,549) | Control<br>(N=4,964) | All<br>(N=6,513) |
| Age (years) | Mean (SD) | 61.7 (14.4) | 69.1 (16.8) | 67.9 (16.6) | 61.7 (14.4) | 63.5 (15.9) | 63.1 (15.6) |
|  | Median (IQR) | 61.7 (52.3, 72.5) | 72.0 (57.7, 82.2) | 70.2 (56.6, 81.0) | 61.6 (52.3, 72.5) | 64.8 (53.4, 75.2) | 63.8 (53.1, 74.7) |
| Age (years) | 18-30 | 33 (2.1%) | 196 (2.5%) | 229 (2.5%) | 33 (2.1%) | 168 (3.4%) | 201 (3.1%) |
|  | 31-40 | 88 (5.7%) | 349 (4.5%) | 437 (4.7%) | 88 (5.7%) | 301 (6.1%) | 389 (6.0%) |
|  | 41-50 | 224 (14.4%) | 634 (8.2%) | 858 (9.2%) | 223 (14.4%) | 554 (11.2%) | 777 (11.9%) |
|  | 51-60 | 388 (25.0%) | 1,151 (14.9%) | 1,539 (16.6%) | 388 (25.0%) | 1,027 (20.7%) | 1,415 (21.7%) |
|  | 61-70 | 385 (24.8%) | 1,345 (17.4%) | 1,730 (18.6%) | 385 (24.9%) | 1,133 (22.8%) | 1,518 (23.3%) |
|  | 71-80 | 278 (17.9%) | 1,887 (24.4%) | 2,165 (23.3%) | 276 (17.8%) | 1,127 (22.7%) | 1,403 (21.5%) |
|  | > 80 | 156 (10.1%) | 2,164 (28.0%) | 2,320 (25.0%) | 156 (10.1%) | 654 (13.2%) | 810 (12.4%) |
| Male |  | 1,005 (64.8%) | 4,264 (55.2%) | 5,269 (56.8%) | 1,005 (64.9%) | 3,037 (61.2%) | 4,042 (62.1%) |
| Ethnicity<sup>(a)</sup> |  | (N=1,336) | (N=6,903) | (N=8,239) | (N=1,336) | (N=4,376) | (N=5,712) |
|  | White | 1,046 (78.3%) | 5,829 (84.4%) | 6,875 (83.4%) | 1,046 (78.3%) | 3506 (80.1%) | 4,552 (79.7%) |
|  | Asian | 153 (11.5%) | 552 (8.0%) | 705 (8.6%) | 153 (11.5%) | 457 (10.4%) | 610 (10.7%) |
|  | Black | 38 (2.8%) | 122 (1.8%) | 160 (1.9%) | 38 (2.8%) | 97 (2.2%) | 135 (2.4%) |
|  | Other | 99 (7.4%) | 400 (5.8%) | 499 (6.1%) | 99 (7.4%) | 316 (7.2%) | 415 (7.3%) |
| Clinically extremely vulnerable status<sup>(b)</sup> |  | (N=1,512) | (N=7,474) | (N=8,986) | (N=1,511) | (N=4,835) | (N=6,346) |
|  | None | 1,411 (93.3%) | 6,722 (89.9%) | 8,133 (90.5%) | 1,410 (93.3%) | 4,460 (92.2%) | 5,870 (92.5%) |
|  | Cancer | 31 (2.1%) | 236 (3.2%) | 267 (3.2%) | 31 (2.1%) | 111 (2.3%) | 142 (2.2%) |
|  | Severe respiratory condition | 27 (1.8%) | 284 (3.8%) | 311 (3.5%) | 27 (1.8%) | 129 (2.7%) | 156 (2.5%) |
|  | Immunosuppression therapy | 35 (2.3%) | 186 (2.5%) | 221 (2.5%) | 35 (2.3%) | 104 (2.2%) | 139 (2.2%) |
|  | Other | 8 (0.5%) | 46 (0.6%) | 54 (0.6%) | 8 (0.5%) | 31 (0.6%) | 39 (0.6%) |
| Comorbidities |  |  |  |  |  |  |  |
|  | None | 472 (30.5%) | 1,974 (25.9%) | 2,446 (26.7%) | 470 (30.5%) | 1,461 (29.7%) | 1,931 (29.9%) |
|  | Diabetes | 386 (24.9%) | 1,826 (23.6%) | 2,212 (23.8%) | 386 (24.9%) | 1,208 (24.3%) | 1,594 (24.5%) |

|  |  |  |  |  |  |  |
| --- | --- | --- | --- | --- | --- | --- |
| **Hypertension** | 584 (37.6%) | 3,188 (41.3%) | 3,772 (40.7%) | 584 (37.7%) | 1,874 (37.8%) | 2,458 (37.7%) |
| **Obesity** | 291 (18.8%) | 865 (11.2%) | 1,156 (12.5%) | 291 (18.8%) | 725 (14.6%) | 1,016 (15.6%) |
| **Chronic Cardiac Disease** | 262 (16.9%) | 2,086 (27.0%) | 2,348 (25.3%) | 262 (16.9%) | 995 (20.0%) | 1,257 (19.3%) |
| **Chronic Pulmonary Disease** | 219 (14.1%) | 1,398 (18.1%) | 1,617 (17.4%) | 218 (14.1%) | 770 (15.5%) | 988 (15.2%) |
| **Asthma** | 261 (16.8%) | 1,091 (14.1%) | 1,352 (14.6%) | 261 (16.8%) | 754 (15.2%) | 1,015 (15.6%) |
| **Other** | 328 (21.1%) | 3,057 (39.6%) | 3,385 (36.5%) | 328 (21.2%) | 1,616 (32.6%) | 1,944 (29.8%) |
| **Number of key comorbidities<sup>(c)</sup>** | **(N=1,546)** | **(N=7,631)** | **(N=9,177)** | **(N=1,542)** | **(N=4,918)** | **(N=6,460)** |
| **0** | 472 (30.5%) | 1,974 (25.9%) | 2,446 (26.7%) | 470 (30.5%) | 1,461 (29.7%) | 1,931 (29.9%) |
| **1** | 471 (30.5%) | 2,486 (32.6%) | 2,957 (32.2%) | 470 (30.5%) | 1,568 (31.9%) | 2,038 (31.5%) |
| **2+** | 603 (39.0%) | 3,171 (41.6%) | 3,774 (41.1%) | 603 (39.1%) | 1,889 (38.4%) | 2,492 (38.6%) |
| **Where COVID was acquired** |  |  |  |  |  |  |
| **Community** | 1,523 (98.1%) | 6,920 (89.6%) | 8,443 (91.0%) | 1,520 (98.1%) | 4,846 (97.6%) | 6,366 (97.7%) |
| **Hospital** | 23 (1.5%) | 704 (9.1%) | 727 (7.8%) | 23 (1.5%) | 94 (1.9%) | 117 (1.8%) |
| **Unknown** | 6 (0.4%) | 102 (1.3%) | 108 (1.2%) | 6 (0.4%) | 24 (0.5%) | 30 (0.5%) |
| **Days since symptom onset (symptomatic patients only):** |  |  |  |  |  |  |
| **Community acquired** | **(N=1,506)** | **(N=6,276)** | **(N=7,782)** | **(N=1,503)** | **(N=4,442)** | **(N=5,945)** |
| Median (IQR) | 7 (4, 9) | 5 (2, 9) | 6 (2, 9) | 7 (4, 9) | 6 (2, 9) | 6 (3, 9) |
| **Hospital acquired** | **(N=11)** | **(N=258)** | **(N=269)** | **(N=11)** | **(N=27)** | **(N=38)** |
| Median (IQR) | 0 (0, 4) | 0 (0, 1) | 0 (0, 1) | 0 (0, 4) | 0 (0, 1) | 0 (0, 1) |
| **Admitted to HDU/ICU at baseline** | **(N=1,526)** | **(N=7,558)** | **(N=9,084)** | **(N=1,524)** | **(N=4,875)** | **(N=6,399)** |
|  | 90 (5.9%) | 145 (1.9%) | 235 (2.6%) | 90 (5.9%) | 136 (2.8%) | 226 (3.5%) |
| **Clinical Frailty Score** | **(N=780)** | **(N=3,965)** | **(N=4,745)** | **(N=779)** | **(N=2,468)** | **(N=4,745)** |
| Median (IQR) | 3 (2, 4) | 4 (2, 6) | 4 (2, 6) | 3 (2, 4) | 3 (2, 5) | 3 (2, 4) |
| **4C Mortality Score** | **(N=1,242)** | **(N=5,443)** | **(N=6,685)** | **(N=1,240)** | **(N=3,692)** | **(N=4,932)** |
| Median (IQR) | 9 (6, 12) | 10 (7, 12) | 10 (7, 12) | 9 (6, 12) | 9 (6, 12) | 9 (6, 12) |
| **Oxygen Saturation (%)** | **(N=1549)** | **(N=7716)** | **(N=9265)** | **(N=1546)** | **(N=4959)** | **(N=6505)** |
| Median (IQR) | 92 (89, 94) | 94 (91, 95) | 93 (90, 95) | 92 (89, 94) | 94 (91, 95) | 93 (90, 95) |
| **Respiratory Rate (Breaths per minute)** |  |  |  |  |  |  |
| Median (IQR) | 24 (21, 28) | 22 (20, 26) | 22 (20, 27) | 24 (21, 28) | 22 (20, 27) | 23 (20, 28) |
| **Glasgow Coma Score: n (%)** | **(N=1,511)** | **(N=7,510)** | **(N=9,021)** | **(N=1,508)** | **(N=4,846)** | **(N=6,354)** |
| Equal to 15 | 1,462 (96.8%) | 6,826 (90.9%) | 8,288 (91.9%) | 1,459 (96.8%) | 4,489 (92.6%) | 5,948 (93.6%) |
| **Urea (mmol/L)** | **(N=1,350)** | **(N=6,192)** | **(N=7,542)** | **(N=1,348)** | **(N=4,127)** | **(N=5,475)** |
| Median (IQR) | 5.7 (4.3, 7.7) | 6.1 (4.5, 8.6) | 6.0 (4.4, 8.5) | 5.7 (4.3, 7.7) | 5.7 (4.2, 7.9) | 5.7 (4.2, 7.9) |
| **C-Reactive Protein (mg/L)** | **(N=1,365)** | **(N=6,104)** | **(N=7,469)** | **(N=1,362)** | **(N=4,102)** | **(N=5,464)** |
| Median (IQR) | 106 (61, 172) | 72 (30, 134) | 79 (34, 140) | 106 (61, 172) | 79 (34, 140) | 85 (40, 148) |

(a) Defined to be mutually exclusive: Asian = any ticked from Arab, East/South/West Asian; Black = Black ticked, but no Asian categories; White = 'White ticked', but not any Asian or Black categories; Other = not categorised Asian, Black, or White, but at least one category ticked in any other box except 'Unknown'.
(b) Categories are not mutually exclusive. No clinically vulnerabilities: all options ticked 'No'; Clinically vulnerable unknown: no boxes are ticked 'Yes', and any level of missingness.
(c) Key comorbidities: Diabetes, Hypertension, Obesity, CCD, CPD, Asthma.

### Treatments received during hospitalisation post baseline

The remdesivir group were generally more medicated post baseline than controls. They were more likely to have been given dexamethasone (93.9% vs. 61.7%) or antibiotics (89.7% vs 79.8%) during hospitalisation. Use of at least one corticosteroid other than dexamethasone was similar in the two groups (9.4% vs 10.7%). In both groups, the use of antiviral agents other than remdesivir was rare – 3.0% of the control group were known to have received an antiviral agent (Supplementary Table 7). Dexamethasone use was identified as a factor that should be adjusted for in all inferential analyses, given its known efficacy as a therapeutic in treating COVID-19.

### In-hospital complications

A complications CRF was uploaded for 6,190 (95.0%) of the matched cohort (remdesivir: 1,504, control: 4,686). The most prevalent complications were viral pneumonia (61.9%), bacterial pneumonia (11.8%), hyperglycaemia (11.0%), acute renal injury/failure (8.9%), anaemia (8.1%) and acute respiratory distress syndrome (ARDS) (7.2%) (Supplementary Table 8). The remdesivir group had higher recorded viral pneumonia (75.2% vs 57.6%), hyperglycaemia (17.8% vs 8.8%), and ARDS (11.4% vs 5.9%). Liver dysfunction (a secondary outcome) was significantly more common in remdesivir treated cases (8.6% vs 5.4%, adjusted odds ratio (OR): 1.51, 95% CI: 1.18-1.92, p=0.0009). Acute renal injury/failure was not associated with treatment group (0.93, 0.75-1.16, p=0.53). No assessment could be made of relatedness nor severity of observed complications as data for this purpose were not collected.

### Primary Outcome

A total of 140/1,507 (9.3%) remdesivir patients and 565/4,734 (11.9%) controls died within 14 days of baseline. 6,202 patients were included in a logistic regression model (Table 2). The OR of death at 14 days for remdesivir vs controls, adjusted for age, sex, number of comorbidities, dexamethasone use, and viral pneumonia was 0.80 with 95% CI (0.60-1.07), p=0.116. Sensitivity analyses did not change inference (see Supplementary Material).

**Table 2:** Analysis of binary/ordinal outcomes.

| Outcome | Incidence | Number included in model | Events | Adjusted Odds-ratio<sup>(a)</sup> | 95% CI | p-value |
| --- | --- | --- | --- | --- | --- | --- |
| **14-day mortality** |  |  |  |  |  |  |
| Remdesivir | 9.3% (140/1,502) | 1,499 | 139 | 0.80 | (0.60, 1.07) | 0.116 |
| Control | 12.0% (565/4,728) | 4,693 | 561 | 1 | - |  |
| **28-day mortality** |  |  |  |  |  |  |
| Remdesivir | 13.6% (203/1,491) | 1,488 | 201 | 0.77 | (0.60, 0.98) | 0.034 |
| Control | 16.6% (777/4,676) | 4,641 | 773 | 1 | - |  |
| **Clinical status at day 15 (+/- 2 days)<sup>(b)</sup>** |  |  |  |  |  |  |
| Remdesivir | - | 1,496 | - | 0.90 | (0.78, 1.06) | 0.230 |
| Control | - | 4,689 | - | 1 | - |  |
| **Use of any non-invasive ventilation during hospitalisation post baseline** |  |  |  |  |  |  |
| Remdesivir | 28.8% (431/1,496) | 1,496 | 437 | 2.7 | (2.18, 3.35) | <0.0001 |
| Control | 10.2% (466/4,587) | 4,564 | 474 | 1 | - |  |
| **Use of any mechanical ventilation or ECMO during hospitalisation post baseline** |  |  |  |  |  |  |
| Remdesivir | 6.8% (106/1,498) | 1,497 | 109 | 1.72 | (1.20, 2.45) | 0.003 |
| Control | 3.3% (153/4,602) | 4,576 | 160 | 1 | - |  |
| **Any acute renal injury or acute renal failure during hospitalisation post baseline** |  |  |  |  |  |  |
| Remdesivir | 8.3% (129/1,503) | 1,499 | 129 | 0.91 | (0.68, 1.20) | 0.487 |
| Control | 8.6% (427/4,964) | 4,637 | 425 | 1 | - |  |
| **Any liver dysfunction during hospitalisation post baseline** |  |  |  |  |  |  |
| Remdesivir | 8.6% (124/1,445) | 1,443 | 124 | 1.51 | (1.19, 1.92) | 0.0008 |
| Control | 5.4% (242/4,504) | 4,483 | 241 | 1 | - |  |

(a) Each odds ratio presented is estimated from a separate model, each adjusted for: baseline age-group (<50, 50-69, 60-69, 70-79, 80+), sex, number of key comorbidities (0, 1, 2+) [where key comorbidities are: any diabetes, hypertension, obesity, CCD, CPD, asthma], dexamethasone use during hospitalisation (Yes, No, Unknown), Viral Pneumonia recorded in the complications CRF (Yes, No, Unknown). Covariate parameter estimates are not published here, as these are not the subject of this study.
(b) Proportional odds-ratio presented – odds of being classified in a worse clinical status at day 15 for Remdesivir compared with Control.

### Secondary Outcomes

Time-to-recovery was found to vary with time and treatment (Figure 3A). During days 1-5, recovery was more likely in controls; during days 6-8 there was no treatment effect; and for days 9-28, remdesivir was associated with a faster recovery (Table 3). Median (IQR) time to recovery was 9 (9-10) days for remdesivir, and 8 (8-9) days for controls. There is evidence to suggest that reduction in 28-day mortality is associated with remdesivir (Table 2): the p-value is 0.03, but uncorrected for multiplicity; this can be interpreted as an estimated 36 (95% CI: 20-290) patients needed to treat to prevent one death. Time-to-death over these 28 days was not significantly associated with treatment group (Figure 3B, Table 3)). Five clinical status classifications were derivable at day 15 (see Supplementary Material). Ordinal regression of this outcome indicates no evidence of an association with treatment group (Table 2). The proportional odds assumption was checked and found to hold. Overall, by Day 15 most patients had improved clinical status compared with baseline: 1,106/1,549 (71.4%) remdesivir vs. 3,303/4,964 (66.5%) controls. Non-invasive ventilation was more likely in the remdesivir group (Table 2). Remdesivir patients were more likely to require invasive mechanical ventilation or ECMO; and where data were available, median (IQR) duration of these interventions in days were 10 (5,16) with remdesivir vs. 6 (3,14) controls. Two outcomes could not be derived due to insufficient daily CRF data: length of time requiring supplementary oxygen, and time-to-first ventilation.

**Figure 3:**
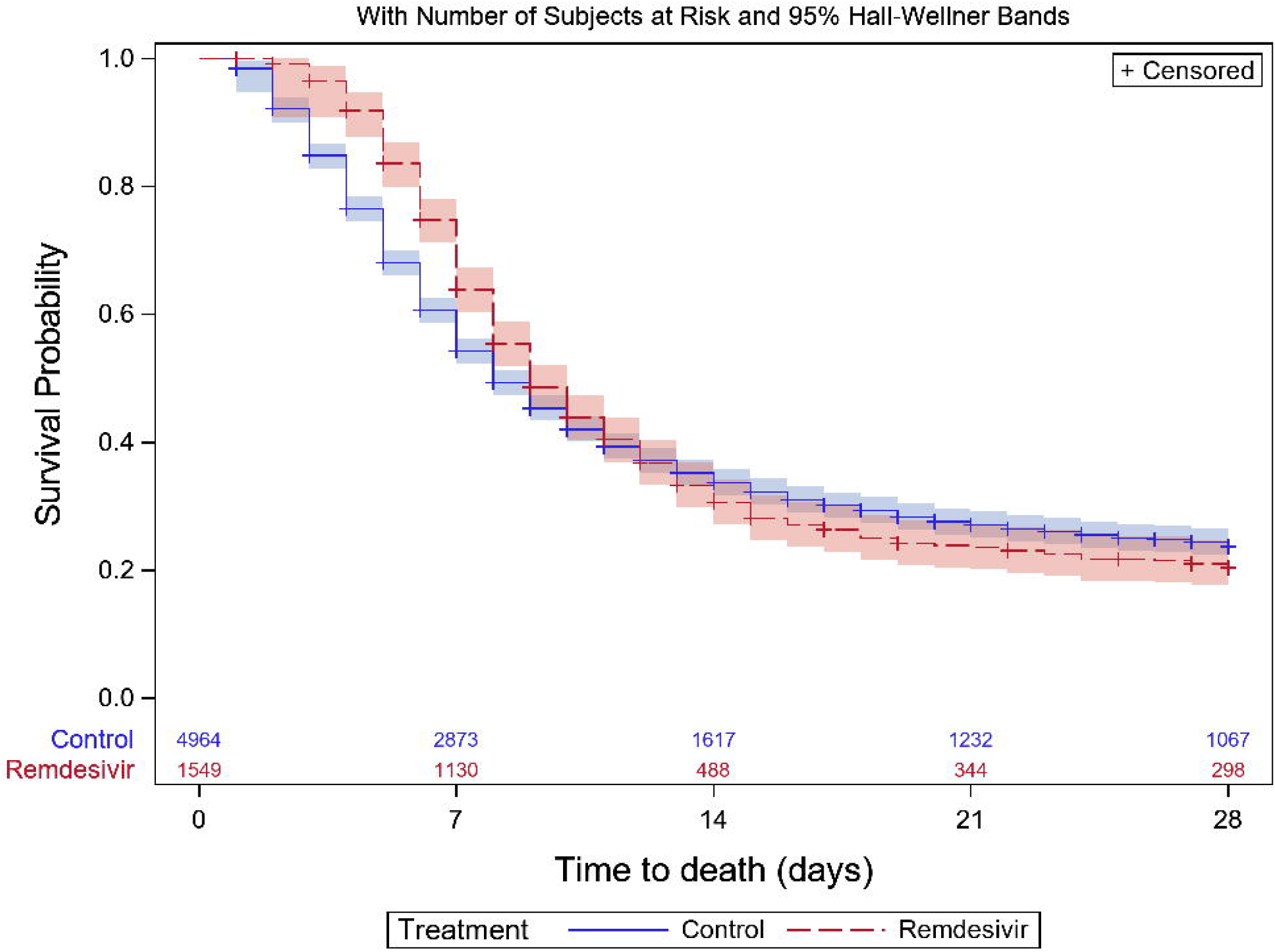
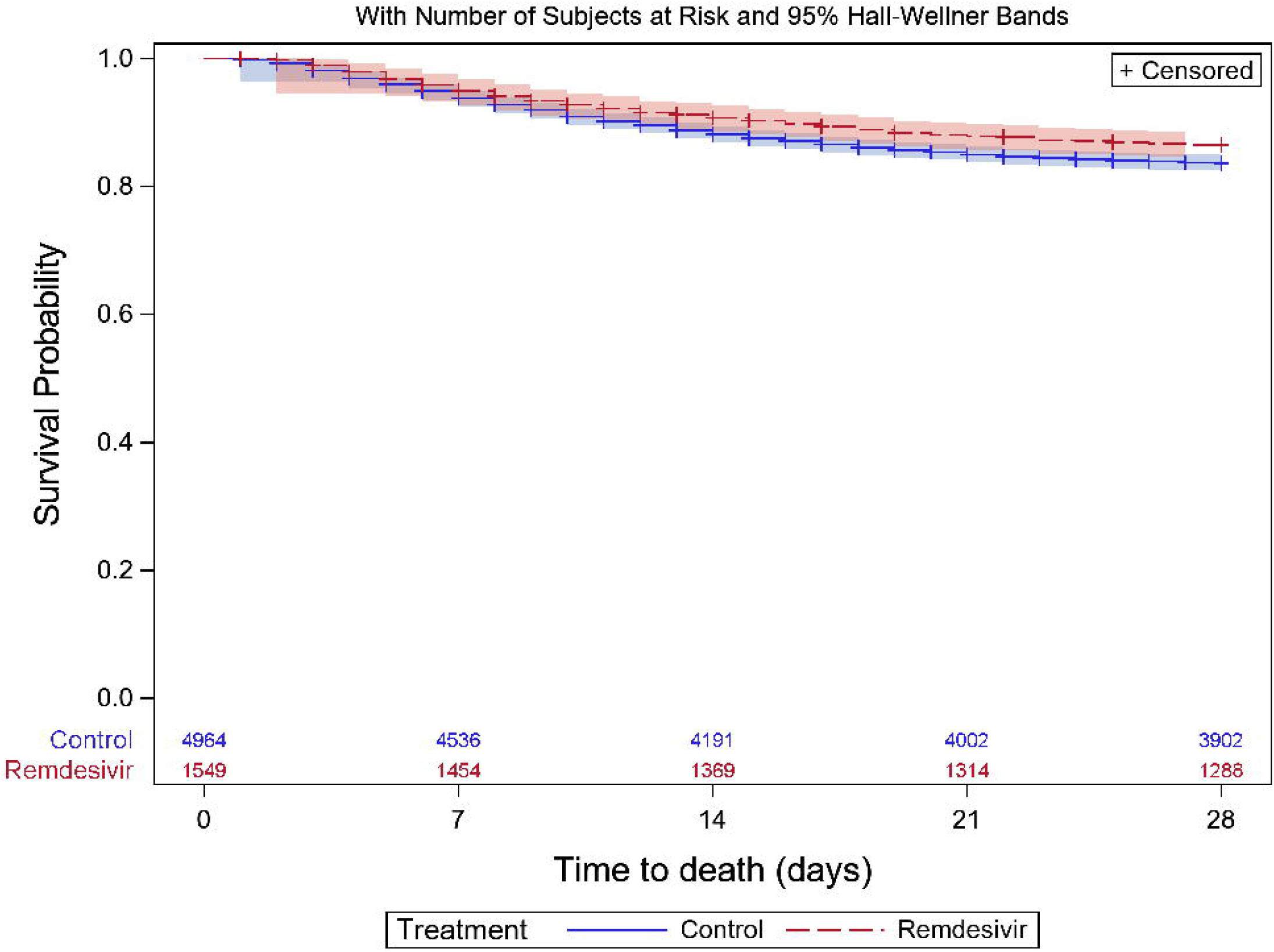
Kaplan-Meier curves showing (A) time-to-recovery, and (B) time-to-death, during first 28 days post baseline, by treatment group

**Table 3:** Analysis of time-to-event outcomes.

| Outcome |  |  | Number included in model | Events | Adjusted Hazard-ratio<sup>(a)</sup> | 95% CI | p-value |
| --- | --- | --- | --- | --- | --- | --- | --- |
| **Time-to-recovery** |  |  |  |  |  |  |  |
|  | **Time period<sup>(b)</sup>:** |  |  |  |  |  |  |
|  | **1-5 days** | Remdesivir | 1543 | 250 | 0.51 | (0.43, 0.60) | <0.0001 |
|  |  | Control | 4918 | 1544 | 1 | - |  |
|  | **6-8 days** | Remdesivir | 1259 | 428 | 1.04 | (0.89, 1.22) | 0.600 |
|  |  | Control | 3184 | 881 | 1 | - |  |
|  | **9-28 days** | Remdesivir | 827 | 517 | 1.22 | (1.06, 1.41) | 0.007 |
|  |  | Control | 2269 | 1159 | 1 | - |  |
| **Time-to-death** |  |  |  |  |  |  |  |
|  |  | Remdesivir | 1,488 | 202 | 0.83 | (0.67, 1.02) | 0.070 |
|  |  | Control | 4,641 | 773 | 1 | - |  |

(a) Each hazard ratio presented is estimated from a separate model, each adjusted for: baseline age-group (<50, 50-69, 60-69, 70-79, 80+), sex, no. of key comorbidities (0, 1, 2+) [where key comorbidities are: any diabetes, hypertension, obesity, CCD, CPD, asthma], dexamethasone use during hospitalisation (Yes, No, Unknown), Viral Pneumonia recorded in the complications CRF (Yes, No, Unknown). Covariate parameter estimates are not published here, as these are not the subject of this study.
(b) Time intervals are chosen such that approximately one third of recoveries took place in each interval. The sample size decreases with each subsequent time-period, as those who recover or are censored prior to the time-period are excluded from the model.

## Discussion

In this analysis of data from a large UK-wide study, we found that remdesivir use was not statistically significantly associated with improved 14-day mortality, when it is given within 24h of hospitalisation due to severe COVID-19, and at a similar time-point, clinical status was not significantly different. The reduction in 28-day mortality estimated in our data was very close to the meta-analysis results published for non-ventilated patients, indicating that our data support current clinical trial findings. We also found that remdesivir patients were more likely to be ventilated and liver dysfunction occurred more frequently. The confidence intervals presented for adjusted odds-ratios indicated considerable uncertainty in our inference. Sensitivity analyses indicated that in the primary outcome at least, this was more likely due to heterogeneity in the population. Study of treatment effects within subgroups would require a larger study.

Our study population was representative of severe COVID-19 patients of all ages, treated in hospitals across the whole UK, predominantly of white ethnic background, and male. Compared with both the ACTT-1 and SOLIDARITY trials, our cohort was older, and less ethnically diverse. As age is a key factor in risk of death from COVID-19 absolute survival outcomes presented here should not be compared directly with the rates published in these trials. Patients in our study received many types of treatment during hospitalisation, but the data do not indicate when these treatments were received, nor the dose, nor duration. Use of other antivirals was rare -the control group can be considered a ‘non-anti-viral therapeutics’ group. There is an indication that dexamethasone use (and by implication corticosteroid use) was greater in the remdesivir group, and use of antibiotics. Complications during hospitalisation indicated some imbalance in the groups. The remdesivir group had higher recorded prevalence of viral pneumonia. This measure is difficult to interpret, and arguably our inclusion criteria define patients that were presenting with viral pneumonia at baseline. It may have little meaning recorded in the complication CRF or may have been a proxy for greater baseline severity, or a secondary effect to the antiviral, though there is less biological plausibility for the latter. Hyperglycaemia, ARDS, and liver dysfunction were also more observed in the remdesivir group. One in eleven patients in our cohort were recorded as suffering from acute renal injury or failure, but this was similar in the two groups. Use of non-invasive ventilation was more likely in the remdesivir group. This could represent a higher level of illness in this group, which was not apparent from the baseline data; alternatively, it could represent a lower threshold for escalation of care in this group, or a perception that escalation was less likely to be futile in this group. A limitation is that we are not able to pinpoint when this ventilation took place, nor for how long it was needed. See also Supplementary Material for further discussion regarding secondary outcomes.

This analysis used data from a prospective observational study, using routine care data collected during a pandemic. There are limitations, in that effectiveness estimates are not from randomised patients, and the data collected reflect local practice by the clinical teams at numerous hospital sites. The study was designed pragmatically to be simple enough to be rapidly implemented, using data that were being collected under a generic protocol. Data completeness for baseline characteristics and final clinical outcomes were found to be extremely good; but daily follow-up data were less available than expected, meaning that two outcomes and clinical status at day 15 could not be derived as planned. Our design created a clear analysis cohort with similar baseline level of severity of COVID-19. Of eligible patients given remdesivir, 43% were excluded because their treatment did not start within 24h of baseline - these patients represent a wider population of treated patients, beyond the scope of our study. Propensity matching was used effectively to select a control group that had a similar profile to remdesivir patients and balanced according to the risk-score diagnostic. The control group were slightly older, but had similar clinical frailty scores, and numbers of comorbidities.

We note that liver dysfunction was increased in the remdesivir group. Raised transaminases are an expected adverse event in nucleoside analogues, and indeed alanine aminotransferase (ALT) was elevated in 7% of remdesivir clinical trial participants [26]. The current data does not allow us to distinguish the level of severity of this liver dysfunction or whether it was reversible.

It is possible that further benefit could be gained if remdesivir, or a similar orally available direct acting antiviral, could be given earlier in the disease process, when pharyngeal shedding and by inference viral replication in the lower respiratory tract is at its highest [12]. SARS-CoV-2-infected rhesus macaques were successfully treated when remdesivir dosing was initiated 12 h after virus inoculum. The authors noted that the efficacy of such direct acting antivirals against acute viral respiratory infections usually drops with time after infection and stressed the importance of dosing humans as quickly as possible. The ACTT-1 trial confirmed that benefits associated with remdesivir were larger earlier in the disease course (<=10 days vs >10 days). Our study contains too few hospital-acquired patients to explore this hypothesis.

Overall, this study in a real-world setting, does not support the findings of the ACTT-1 trial that remdesivir significantly reduces mortality at 14 days in patients hospitalised with severe COVID-19. The results do support meta-analysis evidence that it may provide a reduction in mortality at 28 days. We calculate that the number needed to treat to avoid one death (by 28 days) is 36, where patients do not initially require ventilation. It is possible that this antiviral drug, which shows such promise *in vitro* is being administered too late in the disease process to have its optimum impact.

## Data Availability

This work uses data provided by patients and collected by the NHS as part of their care and support #DataSavesLives. ISARIC4C welcomes applications for data and material access through our Independent Data and Material Access Committee (https://isaric4c.net).

## Acknowledgments

Dr Emily Granger of the London School of Hygiene and Tropical Medicine, for advice regarding optimal balance diagnostics. Dr Lance Turtle of the University of Liverpool, for insight into clinical decision making during the treatment of COVID-19 patients in hospital.

## Authorship

Conceptualisation: BN Arch, A Rosala-Hallas, EM Harrison, AP Jones, MG Semple. Formal analysis: BN Arch, A Rosala-Hallas. Writing original draft: BN Arch, D Kovacs, JT Scott. Writing reviewing and editing: BN Arch, JK Baillie, CG Gamble, EM Harrison, AP Jones, PJM Openshaw, JT Scott, MG Semple.

## Ethical approval

Ethical approval for data collection was given by the South Central - Oxford C Research Ethics Committee in England (Ref: 13/SC/0149) and by the Scotland A Research Ethics Committee (Ref: 20/SS/0028).

## Declaration of interests

All authors have completed the ICMJE uniform disclosure form at www.icmje.org/coi_disclosure.pdf and declare:

BNA, ARH, APJ and CGG report: the manufacturer of remdesivir, Gilead, is involved in funding trials that the Liverpool Clinical Trials unit is co-ordinating: a randomised controlled trial (HART-CT) that is fully funded by Gilead and sponsored by the University of Liverpool; and a trial (RIAltO) that is part funded by Gilead. APJ is the lead statistician on the HART-CT trial. PJMO reports personal fees from consultancies and from the European Respiratory Society; grants from the Medical Research Council (MRC), MRC Global Challenge Research Fund, EU, NIHR BRC, MRC/GSK, Wellcome Trust, NIHR (Health Protection Research Unit [HPRU] in Respiratory Infection); and is an NIHR senior investigator outside of the submitted work; his role as President of the British Society for Immunology was unpaid but travel and accommodation at some meetings was provided by the Society. MGS reports grants from NIHR UK, MRC UK, and HPRU in Emerging and Zoonotic Infections, University of Liverpool during the conduct of the study.

## Funding

ISARIC4C is funded by two major awards from the Medical Research Council (MRC; grant MC_PC_19059), and The National Institute For Health Research (NIHR; award CO-CIN-01). PJMO is supported by a NIHR Senior Investigator Award [award 201385]. The Liverpool clinical trials unit did not receive any direct funding for this work. DK is funded by UK MRC Precision Medicine Training Grant (MR/N013166/1-LGH/MS/MED2525). The views expressed are those of the authors and not necessarily those of the UKRI, NHIR, or MRC.

## Notes

### Clinical Protocols

https://isaric4c.net/protocols

### Summary of Updates

One year on from conducting our study, we have updated the paper slightly to reflect it's current relevance as adding to the evidence base rather than presenting novel evidence. It is also updated to incorporate further results published from the WHO SOLIDARITY trial.

