## Supplementary Material for "Evaluation of the effectiveness of remdesivir in severe COVID-19 using observational data from a prospective national cohort study"

### 2. Inclusion and Exclusion Criteria

Include patients that satisfy all the following inclusion criteria:

- Laboratory confirmed SARS-CoV-2 infection
- Hospitalised at baseline
- Aged ≥ 18 years[1] at baseline
- Requiring supplementary oxygen[2] ($SpO_2$ ≤ 94%) at any time during 24 hours post baseline
- RDV initiated ≤ 24 hours post baseline (RDV group only)[3]

Exclude patients for whom any of the following are confirmed:

- Requiring a high flow cannula, any ventilation or ECMO during first 24 hours post baseline[4]
- Pregnant
- Chronic kidney disease[5]
- Is a COVID-19 re-admission (was previously hospitalised for COVID-19)
- Missing Day 1 Daily Treatment CRF[6]
- Missing 'Medication' section of the Outcome CRF[7]
- RDV initiated > 24 hours post baseline

---

1 RDV was recommended in people aged 12 and over and weighing over 40kg. Weight was not generally measured at baseline for practical reasons, so we have excluded all children to be sure that no-one in the control group was likely to weigh less than 40kg.
2 We assessed this using the Admission CRF and the Day 1 Daily CRF – a patient was judged as requiring oxygen if it was confirmed that they were given oxygen in either of these CRFs, or if $SpO_2$ ≤ 94%
3 If RDV was given, we required this to have been confirmed starting within 1 day of baseline – this is because data collection of outcomes is related to baseline. Whilst RDV may not be always given straight away, this condition makes it easier to obtain two clear groups for the purpose of comparison.
4 We assessed this using the Admission CRF and the Day 1 Daily CRF – any evidence within these of high-flow cannula, or any ventilation in either of these CRFs led to patients being excluded
5 This was a comorbidity that could be reported at baseline. Patients were also deemed as potentially suffering from CKD if their baseline eGFR was <30.
6 A missing Day 1 Daily CRF means that baseline is unknown. A missing treatment section from this form makes it difficult to assess oxygenation or ventilation needs.
7 This is required to confirm whether patients received any RDV during hospitalisation, and the dates of the first and last doses.

### 3. Rationale For and Definitions of Outcomes

These are summarised in Supplementary Table 1. Outcomes were chosen to mirror clinical trial reporting and be potentially measurable with the observational data that were being collected. 14-day mortality was chosen as the primary outcome as there was a benefit for remdesivir observed in the ACTT-1 trial [1] for this outcome, and as this was not reported by SOLIDARITY [2]. Mortality data also has the greatest potential for accuracy in an observational study. Length of hospital stay was a key outcome of the SOLIDARITY trial but is not listed as an outcome here. This is because our cohort includes hospital acquired COVID patients, and some patients with long-term institutionalisation. We are more interested in COVID-related hospital stay, but data on this are not collected beyond 28 days, and so we use the outcome 'Time-to-recovery' to double as inference to test whether remdesivir impacts on short-term length of stay.

For all outcomes, Day 1 was the date of baseline. The primary outcome was 14-day mortality - deaths were counted if they occurred on or before Day 14. 'Time-to-recovery' was defined as the number of days from baseline to discharge from hospital or to cessation of COVID-19 related healthcare (whichever occurred first). All other patients were censored at last available follow-up or 28 days (whichever occurred first), and deaths before 28 days, with no prior recovery, were censored at day 28 (in line with the definition of this outcome in the ACTT-1 trial). '28-day mortality' was defined in the same way as 14-day mortality. 'Time-to-death' was defined as the number of days from baseline to date of death. All other patients were censored at last available follow-up or 28 days (whichever occurred first). 'Clinical status at day 15' was defined in the same way as for the ACTT-1 trial but with potential limitations on its derivation due to the observational nature of our data. 'Use of any non-invasive ventilation' (NIV) during hospitalisation was a binary outcome recorded in the Outcome CRF, and daily use of NIV is also recorded in daily CRFs where these existed.' Use of any mechanical ventilation or ECMO' was a binary outcome derived from positive indicators in the Outcome CRF of any (a) invasive ventilation, (b) any inhaled nitric oxide, (c) tracheostomy inserted, or (d) ECMO support. As for NIV, daily CRFs were also checked for positive indicators of any of these interventions. Incidence of acute renal injury/failure and incidence of liver dysfunction were binary outcomes derived from specific indicators of these complications in the Outcome CRF, and where these complications were mentioned in the free-text field provided for the description of 'Other complications'.

**Supplementary Table 1: Outcome definitions and rationale**

| Outcome | Definition | Rationale |
|---|---|---|
| 14-day mortality | Death from any cause during day 1 to day 14 inclusive, where day 1 is baseline | In the ACTT-1 trial, there was a significant benefit over placebo for this outcome in the patients at clinical status*=5 at baseline. This outcome was not reported on in the SOLIDARITY interim report. |
| Time-to-recovery | Recovery is defined as discharge from hospital or continued hospitalisation with no on-going health-care needs related to COVID-19. For patients that recover, a time-to-event is defined as the number of days from baseline to discharge, or from baseline to cessation of COVID-19 related healthcare (whichever occurs first). All other patients are censored at last available follow-up or 28 days (whichever occurs first). All deaths before 28 days (with no prior recovery) are censored at day 28, (this is to mirror the definition for this outcome in the ACTT-1 trial). | This is the primary outcome of the ACTT-1 trial, and a significant benefit for remdesivir was reported. This outcome was not significant in the SOLIDARITY trial. |
| 28-day mortality | Death from any cause during day 1 to day 28 inclusive, where day 1 is baseline | A significant benefit was found for this outcome in the ACTT-1 trial, but not in SOLIDARITY. |
| Time-to-death | We restrict this analysis to deaths of any cause within 28 days of baseline. Any patient alive at the end of the study period was censored at 28 days. Any patient that left hospital alive prior to 28-days is assumed alive at 28 days unless data from the final outcome CRF indicated a death during that time. Any patient lost to follow-up prior to 28 days was censored at their last known follow-up. | A significant benefit of remdesivir was found for this outcome in the ACTT-1 trial. This outcome was not reported on in the SOLIDARITY interim report. |
| Clinical status at day 15 | Clinical status is defined as an 8-point ordinal score with the following categories: (1) Not hospitalised, and not limited physically; (2) Not hospitalised, but limited physically and/or requiring supplementary oxygen; (3) Hospitalised but medically OK; (4) Hospitalised, not requiring supplementary oxygen, but needs on-going COVID related care; (5) Hospitalised and requiring supplementary oxygen; (6) Hospitalised and requiring non-invasive ventilation; (7) Hospitalised and requiring mechanical ventilation or ECMO; (8) Dead. This was to mirror the definition for this outcome in the ACTT-1 trial. | A significant benefit of remdesivir was found for this outcome in the ACTT-1 trial. This outcome was not reported on in the SOLIDARITY interim report. |
| Length of time receiving supplementary oxygen | Number of days a patient is recorded as having been receiving supplementary oxygen. On days where there is no daily CRF, a planned method of imputation is defined (see Statistical Analysis Plan). | This is a secondary outcome of the ACTT-1 trial. |
| Time to first ventilation | The Daily CRF was to be used to define, where possible, date of first use of ventilation. Any patient not recorded as ventilated that was lost to follow-up or died before 28 days, was to be censored at the date of last follow-up, or date of death. | This was not an outcome of the clinical trials, but of interest to the designers of this study. |

| | | |
|---|---|---|
| Use of non-invasive ventilation at any time during 28 days post baseline | This is a binary outcome indicating whether there was any use of non-invasive ventilation during hospitalisation. This is recorded in the Interim Outcome CRF. Daily CRFs were also checked for any positive indicators of the outcome. | This was not an outcome of the clinical trials, but measurable in this study. ACTT-1 measured duration of non-invasive ventilation. |
| Use of mechanical ventilation / ECMO at any time during 28 days post baseline | This is a binary outcome indicating whether there was any use of invasive mechanical ventilation or ECMO during hospitalisation. This is recorded in the Interim Outcome CRF and is derived from positive indicators of any (a) invasive ventilation, (b) any inhaled nitic oxide, (c) tracheostomy inserted, or (d) Extracorporeal (ECMO) support. Daily CRFs were also checked for any positive indicators of the outcome. | This was not an outcome of the clinical trials, but measurable in this study. ACTT-1 measured duration of invasive mechanical ventilation or ECMO. |
| Acute renal injury/acute renal failure at any time during 28 days post baseline | This is a binary outcome indicating whether there was any observed acute renal injury or failure during hospitalisation. This is recorded in the Complications section of the Interim Outcome CRF. | This was not an outcome of the clinical trials, but measurable in this study. |
| Liver dysfunction at any time during 28 days post baseline | This is a binary outcome indicating whether there was any observed liver dysfunction during hospitalisation. This is recorded in the Complications section of the Interim Outcome CRF. | This was not an outcome of the clinical trials, but measurable in this study. |

*See definition for Clinical Status at day 15

### 4. Policy Guidelines for use of Remdesivir: Evolution over time

Patients were recommended to receive 200mg on Day 1 of treatment, followed by 100mg daily for 4 days. An optional additional 5 days of 100mg daily was recommended till 29th September, but the wording changed over time:

26th May to 2nd June: "If a patient does not demonstrate clinical improvement"

3rd June to 28th September: "If a patient deteriorates and progresses to ventilation and/or ECMO treatment"

29th September to 11th November: "Ensure that clinicians prescribe a maximum treatment course of 5 days" – this was due to a disruption in supply of the drug. It is stated that some exceptions may receive additional doses, but these were undefined by the guidelines.

12th November onwards: "The use of remdesivir should be reassessed daily. Consider stopping remdesivir if: (a) the patient clinically improves and no longer requires supplemental oxygen 72 hours after commencement of treatment; or (b) the patient continues to deteriorate despite 48 hours of sustained mechanical ventilation."

### 5. Additional information regarding data collection and case report forms

The Chief Medical Officer (CMO) for Her Majesty's Government UK mandated that data on all patients that receive any drugs through the EAMS must be captured through the International Severe Acute Respiratory and Emerging Infection Consortium (ISARIC) WHO Clinical Characterisation Protocol UK (CCP-UK) case report forms (CRFs) [3]. Completion of the mandatory fields of the CCP-UK CRFs is required, to support ongoing implementation, including determination of future drug allocations between NHS hospitals, and evaluation. The CMO commissioned this analysis of effectiveness and safety based on the pragmatic data being collected by CO-CIN.

**Complications data:**

It is important to recognise that in an observational study, a complication is an event associated with the admission for the disease under observation and not necessarily a result of exposure to treatment. The ISARIC WHO CCP case report form is designed for generic disease characterisation of emerging or novel pathogens, so prior assumptions are deliberately not made whether to define disease features as a complication that is primary to the disease characteristic (severe spectrum) or secondary to the disease process.

**Treatment and intervention data:**

As this study is observational, patients were treated according to local standard of care. Some treatment data are collected in daily CRFs, but these are not routinely completed for all patients, and certainly not every day. Detailed data on remdesivir use were collected by design, but other treatments are restricted to a general assessment in the Outcome CRF. This goes for medication as well as ventilation methods and ICU/HDU.

### 6. Reasons for exclusion

The eligible cohort was defined to be the sub-population of patients most likely to benefit from receiving remdesivir. 28,896 (73%) of UK patients hospitalised with COVID-19 between 26th May and 30th November 2021 were not in this cohort (see Supplementary Table 2). Of those within the eligible cohort (n=10,434), 2,707 (26%) received remdesivir at some point during the first 28 days post baseline. 1552 (57%) of these had remdesivir initiated within 24 hrs of baseline – these patients form the treatment group of our study. The remaining 1,155 patients form a third potential treatment group but are excluded from our study as the dataset does not enable us to define their clinical status at initiation of remdesivir. Timing of initiation is summarised in Supplementary Table 3. Their baseline characteristics and outcomes are described in Section 7 below.

**Supplementary Table 2: Summary of baseline factors excluding patients from the full cohort (these are not mutually exclusive)**

| Reason for exclusion | Total (N=39,330) |
|---|---|
| SARS-CoV-2 infection not confirmed | 646 (1.6%) |
| Not confirmed hospitalised at baseline (hospital admission date after Day 1 CRF date), or hospital admission date missing) | 338 (0.9%) |
| Aged < 18 years at baseline | 769 (2.0%) |
| Did not require oxygen at baseline / could not be ascertained | 20,517 (52.2%) |
| Patient required a high-flow cannula, any ventilation or ECMO during first 24 hours post baseline | 4,657 (11.8%) |
| Patient was pregnant | 667 (1.7%) |
| Evidence of Chronic kidney disease (CKD ticked, or baseline eGFR <30) | 7,182 (18.3%) |
| COVID-19 re-admission | 1,640 (4.2%) |
| No Treatment CRF | 1,631 (4.1%) |

**Supplementary Table 3: Remdesivir patients within eligible cohort, but excluded as initiation was not confirmed as within 24h of baseline**

| | |
|---|---|
| Remdesivir not confirmed as within 24 hours of baseline | 1,155 (43.3%) |
| Remdesivir initiated: | |
| On days 2 or 3 | 746 (64.5%) |
| On days 4, 5,6, or 7 | 284 (24.6%) |
| After day 8 (maximum day = 28) | 77 (6.7%) |
| Date Unknown | 16 (1.4%) |
| Date of initiation entered as prior to baseline – likely data entry error | 32 (2.8%) |

### 7. Remdesivir patients within eligible cohort with initiation of treatment > 24hrs post baseline, or unknown

A subgroup of the eligible cohort was excluded from our analysis, as they received remdesivir but not within 24 hours of baseline. Their baseline characteristics are broadly similar to the remdesivir patients included in our analysis, though on average approximately 3-5 years older, less likely to have a clinical vulnerability but more likely to have at least one key comorbidity (Supplementary Table 4).

**Supplementary Table 4: Baseline characteristics of patients in the eligible cohort that received any remdesivir**

| | Remdesivir not initiated at baseline (N=1,155) | Remdesivir initiated at baseline (N=1,552) |
|---|---|---|
| **Age (years)** | | |
| Mean (SD) | 65.4 (15.1) | 61.7 (14.4) |
| Median (IQR) | 66.3 (55.7, 76.9) | 61.7 (52.3, 72.5) |
| **Age (years)** | | |
| **18-30** | 22 (1.9%) | 33 (2.1%) |
| **31-40** | 58 (5.0%) | 88 (5.7%) |
| **41-50** | 119 (10.3%) | 224 (14.4%) |
| **51-60** | 237 (20.52%) | 388 (25.0%) |
| **61-70** | 250 (21.65%) | 385 (24.8%) |
| **71-80** | 289 (25.02%) | 278 (17.9%) |
| **> 80** | 180 (15.6%) | 156 (10.1%) |
| **Male** | 692 (60.0%) | 1,005 (64.8%) |
| **Ethnicity[a]** | **(N=1,023)** | **(N=1,336)** |
| **White** | 817 (79.9%) | 1,046 (78.3%) |
| **Asian** | 103 (10.1%) | 153 (11.5%) |
| **Black** | 34 (3.3%) | 38 (2.8%) |
| **Other** | 69 (6.7%) | 99 (7.4%) |
| **Clinically extremely vulnerable status[b]** | **(N=1,113)** | **(N=1,512)** |
| **None** | 1,010 (87.5%) | 1,411 (93.3%) |
| **Cancer** | 38 (3.3%) | 31 (2.1%) |
| **Severe respiratory condition** | 39 (3.4%) | 27 (1.8%) |
| **Immunosuppression therapy** | 38 (3.3%) | 35 (2.3%) |
| **Other** | 7 (0.6%) | 8 (0.5%) |
| **Comorbidities** | **(N=1,131)** | **(N=1,546)** |
| **None** | 271 (24.0%) | 472 (30.5%) |
| **Diabetes** | 297 (26.3%) | 386 (24.9%) |
| **Hypertension** | 477 (42.2%) | 584 (37.6%) |
| **Obesity** | 208 (18.4%) | 291 (18.8%) |
| **Chronic Cardiac Disease** | 245 (21.7%) | 262 (16.9%) |
| **Chronic Pulmonary Disease** | 211 (18.7%) | 219 (14.1%) |
| **Asthma** | 189 (16.7%) | 261 (16.8%) |
| **Other** | 331 (29.3%) | 328 (21.1%) |
| **Number of key comorbidities[c]** | **(N=1,131)** | **(N=1,546)** |
| **0** | 271 (24.0%) | 472 (30.5%) |
| **1** | 362 (32.0%) | 471 (30.5%) |
| **2+** | 508 (44.9%) | 603 (39.0%) |
| **Admitted to HDU/ICU at baseline** | **(N=1,135)** | **(N=1,526)** |
| | 31 (2.7%) | 90 (5.9%) |
| **Clinical Frailty Score** | **(N=579)** | **(N=780)** |
| Median (IQR) | 3 (2, 5) | 3 (2, 4) |
| **Where COVID was acquired** | | |
| **Community** | 1,101 (95.3%) | 1,523 (98.1%) |
| **Hospital** | 50 (4.3%) | 23 (1.5%) |
| **Unknown** | 5 (0.4%) | 6 (0.4%) |
| **Community acquired** | **(N=1,082)** | **(N=1,506)** |
| Median (IQR) | 6 (3, 9) | 7 (4, 9) |
| **Hospital acquired** | **(N=27)** | **(N=11)** |
| Median (IQR) | 0 (0, 1) | 0 (0, 4) |

### 8. Propensity Score Modelling

#### 8.1. Methods

A logistic regression with dependent variable remdesivir (Yes/No) was fitted including covariates chosen for their likelihood to affect use of remdesivir and/or the primary outcome: month of baseline; ISARIC-4C tier of participating centre (0 or 1, 2); sex; age; broad ethnicity group (White, Asian, Black, Other); clinically extremely vulnerable status (Yes: [Any of the following: cancer, severe respiratory condition, on immunosuppression therapy, other], none, or unknown); binary indictors of the key comorbidities: diabetes, hypertension, obesity, chronic cardiac disease (CCD), chronic pulmonary disease (CPD), and asthma; where COVID was acquired (Community / Hospital); admitted to HDU/ICU at baseline (Y/N). A pragmatic approach to model fitting was taken – the list defined all baseline variables available that were considered potentially related to either the likelihood of receiving remdesivir, or the likelihood of the primary outcome; we planned that it might be necessary for categories to be collapsed to simplified discriminators (e.g., 'White' vs Other ethnicity, or 'Total number of comorbidities'), should the model become over-parametrised. We intended that $2^{nd}$-order interactions could be investigated, and if statistically significant, added to the model. The fitted propensity model was used to calculate the probability of receiving remdesivir for each patient according to their baseline factors. Patients who were given remdesivir were matched to control patients with a similar PS who did not get remdesivir, using a nearest neighbour matching algorithm and a pre-specified maximum difference in propensity score (calliper width of 0.2 standard deviations). The ratio of remdesivir patients to controls was restricted to be at most 1:4 (Rassen *et al* [4] suggest that 1:*n* matching leads to higher precision in estimating effect sizes, with a cost of a small increase in bias with increasing *n*). Controls were chosen without replacement, so could not be matched to more than one remdesivir patient.

The matched cohort was checked for balance, by using the disease risk score diagnostic described by Stuart *et al* [5] (identified as a good overall diagnostic through simulation studies by Granger *et al* [6] *(key findings yet to be published*). An underlying primary outcome risk score was calculated using the following analysis dataset: all non-remdesivir patients satisfying inclusion/exclusion criteria; and modelling 14-day mortality using logistic regression, with key risk factors fitted as covariates: sex, age-group (<50, 50-69, 70-79, 80+), and number of key comorbidities confirmed (0, 1, 2+, from diabetes, hypertension, obesity, CCD, CPD, asthma) (chosen from Knight *et al* [7]). The fitted model was used to calculate a primary outcome risk score (RS) for each patient included in the matched cohort. The groups were checked for balance by calculating the weighted absolute standardised difference in RS using the R package *MatchIt* [8], *specifying* the risk score as an additional variable to be summarised for balance. NB weights in this method are similar to those used in the outcome analysis for this study, but also multiplied by $n_T/n_C$, the ratio of treated to controls in the matched cohort. (Remdesivir patients are given weight 1, and controls are given weight $\frac{1}{k_j}\left(\frac{n_T}{n_C}\right)$, where $k_j$ is the number of controls also matched to the remdesivir patient *j*. We predefined an $ASD_{RS} < 0.1$ as a diagnostic to indicate adequate balance between the matched groups.

If matching was found to have been unsuccessful in balancing treatment and control groups, the propensity score model was revised. Further discussion of assessment of balance is given in 8.3 below.

#### 8.2. Results

Of the 9,278 patients satisfying inclusion and exclusion criteria, a total of 9,265 patients (1,549 RDV, 7,716 controls) were included in the propensity score analysis and could be potentially included in the matched cohort. 13 were excluded, as their sex was either not recorded or not specified. As this is a key matching variable, but the number missing is so small, it was decided to simply exclude these patients from further analysis.

Several propensity score models were tried, using the matching variables pre-defined in the statistical analysis plan (v1.0), and tested for balance. These were:

**M1** All potential matching variables included, with month of baseline and age included as continuous

**M2** As M1, but with age included as a categorical variable using the categorisations of the 4C Mortality Score defined by Knight et al. (2020)

**M3** As M2, but with month of baseline included as a categorical variable

**M4** As M2, but with non-significant covariates removed

**M5** As M4, and adding in any 2-way interactions that are found to be significant

A balance statistic was calculated for each model, and the model that gave the optimal balance between matched groups was chosen: Model 4 (M4). This gave a weighted $ASD_{RS}$ of 0.01. Model results are presented in Supplementary Table 5. Distributions of propensity scores by treatment group in the eligible cohort, and in the matched cohorts are presented in Supplementary Figure 1. The disease risk score model results are presented in Supplementary Table 6.

**Supplementary Table 5: Chosen propensity score model: Logistic regression model fitted to the full analysis cohort (n=9,265) modelling chance of being given remdesivir as a function of baseline characteristics**

| Model term | Parameter Estimate | Standard Deviation | 95% CI | SMD*** |
|---|---|---|---|---|
| (Intercept) | -2.87 | 0.21 | (-3.28, -2.46) | |
| Month of baseline* | 0.17 | 0.02 | (0.13, 0.21) | 0.09 |
| Male | -0.33 | 0.06 | (-0.45, -0.21) | 0.01 |
| Aged 50-59 | 0.21 | 0.09 | (0.03, 0.39) | 0.02 |
| Aged 60-69 | 0.22 | 0.09 | (0.04, 0.40) | 0.02 |
| Aged 70-79 | -0.34 | 0.1 | (-0.54, -0.14) | 0.05 |
| Aged 80+ | -0.88 | 0.11 | (-1.10, -0.66) | 0.02 |
| Asian | 0.15 | 0.1 | (-0.05, 0.35) | 0.01 |
| Black | 0.29 | 0.2 | (-0.10, 0.68) | 0.01 |
| Other | 0.1 | 0.12 | (-0.14, 0.34) | 0.00 |
| Ethnic group unknown | 0.31 | 0.09 | (0.13, 0.49) | 0.00 |
| Clinically vulnerable** | -0.32 | 0.12 | (-0.56, -0.08) | 0.01 |
| Unknown whether clinically vulnerable | -0.09 | 0.16 | (-0.40, 0.22) | 0.00 |
| Obese | 0.37 | 0.08 | (0.21, 0.53) | 0.05 |
| Unknown obesity status | -0.1 | 0.09 | (-0.28, 0.08) | 0.00 |
| Chronic cardiac disease | -0.25 | 0.08 | (-0.41, -0.09) | 0.02 |
| Unknown chronic cardiac disease status | -0.37 | 0.19 | (-0.74, 0.00) | 0.01 |
| Hospital acquired | -1.63 | 0.22 | (-2.06, -1.20) | 0.00 |
| Unknown whether hospital or community acquired | -1.19 | 0.43 | (-2.03, -0.35) | 0.00 |
| Admitted to ICU/HDU at baseline | 0.95 | 0.14 | (0.68, 1.22) | 0.05 |
| Unknown whether admitted to ICU/HDU at baseline | -0.18 | 0.22 | (-0.61, 0.25) | 0.00 |

Reference categories: Female; Aged <50 yrs; White ethnic group; Community acquired COVID; and for all other categories, absence of the characteristic, e.g. No obesity. Parameter estimate are log-odds ratios.

*Month of baseline is fitted as continuous. The parameter estimate represents the average increase in log-odds of receiving Remdesivir with each passing month.

**Clinically vulnerable: if any of the categories were ticked 'Yes' in the 'Member of a clinically extremely vulnerable group' section of the CRF; Not clinically vulnerable: all options ticked 'No'; Clinically vulnerable unknown: no boxes are ticked 'Yes', and any level of missingness.

***Standard mean difference (SMD). This is a post-hoc balance measure output by the package *MatchIt*. These are adjusted for ratio of controls to treated by incorporating weights (see Section 7.1 above). SMD < 0.05 indicates good balance for variables that are prognostic of outcomes; SMD < 0.1 indicates good balance for variables that are not prognostic of outcomes.

**Supplementary Figure 1: Histograms showing the distributions of propensity scores by treatment group: full analysis cohort (Raw), and matched cohort**

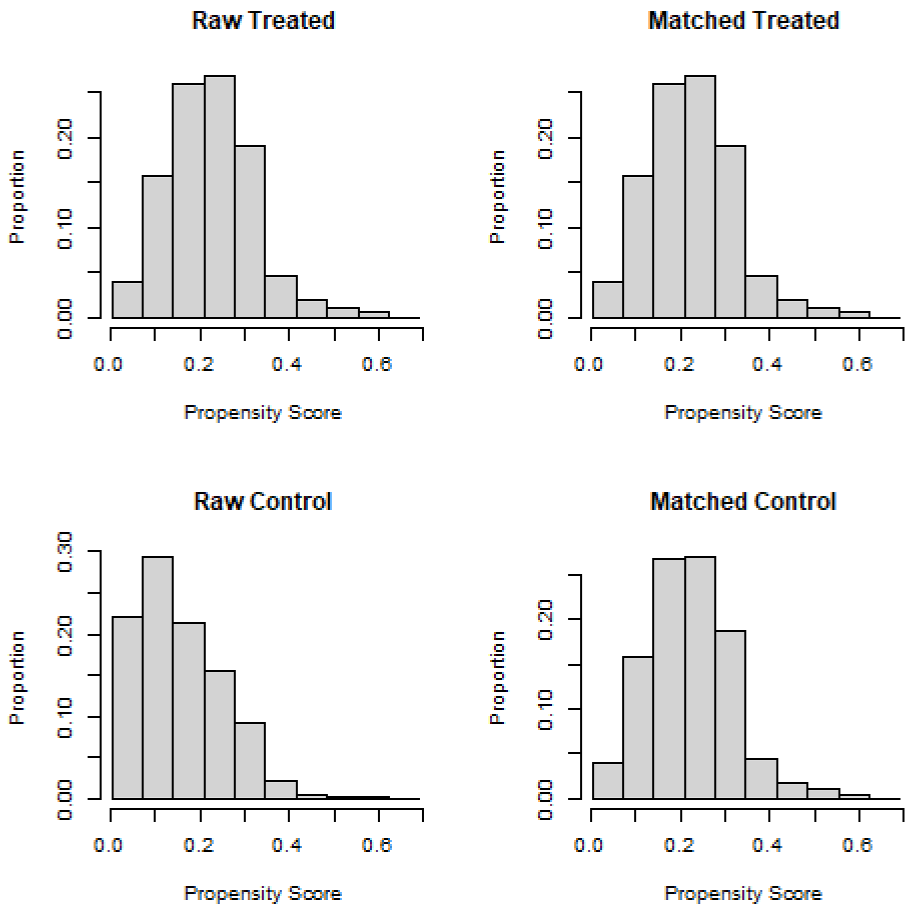

**Supplementary Table 6: Risk score model: Logistic regression modelling 14-day mortality as a function of pre-defined baseline risk factors (n=6,757*)**

| Model term | Parameter Estimate | Standard Deviation | 95% CI |
|---|---|---|---|
| (Intercept) | -4.09 | 0.25 | (-4.58, -3.60) |
| Male | 0.31 | 0.07 | (0.17, 0.45) |
| Aged 50-59 | 0.92 | 0.29 | (0.35, 1.49) |
| Aged 60-69 | 1.61 | 0.27 | (1.08, 2.14) |
| Aged 70-79 | 2.42 | 0.25 | (1.93, 2.91) |
| Aged 80+ | 3.1 | 0.25 | (2.61, 3.59) |
| One key comorbidity** | 0.17 | 0.1 | (-0.03, 0.37) |
| Two or more key comorbidities | 0.42 | 0.1 | (0.22, 0.62) |

Reference categories: Female; Aged <50 yrs; No key comorbidities.

*Patients in the eligible cohort that did not receive any remdesivir, with complete data for the outcome and covariates fitted

**Key comorbidity: Obesity, any diabetes, hypertension, chronic cardiac disease, chronic pulmonary disease, asthma

#### 8.3. Checking for balance at baseline

In this section, additional post-hoc assessments of balance are presented. Where treated patients are matched to a variable number of controls, it is not possible to use a standard Table 1 of baseline statistics to transparently compare groups for balance [4]. We have presented baseline characteristics in this paper, to summarise each cohort before and after matching, rather than to assess balance. Traditional measures of balance for individual baseline variables such as standardised mean differences (SMDs) also cannot be straightforwardly calculated. For the variables included in the propensity score model, weighted SMDs are output as standard from the *summary* function within *MatchIt* [8]. These are presented in Table 3 above. An SMD < 0.0.5 is considered to imply adequate balance where variables are prognostic of outcomes – so although in Table 1 we have potential differences in the cohorts within age-groups, SMDs do not indicate that this is sufficient to incur bias. SMDs for other variables in Table 1 (other than age as a continuous variable) cannot be calculated using *MatchIt*, due to the presence of missing data. Manual calculation of these statistics in the presence of missing data is beyond the scope of this study. Distributions of continuous baseline variables in Table 1 are presented graphically in Supplementary Figure 2. This shows that the groups were well matched for oxygen saturation, respiratory rate, urea and 4C mortality risk score. When age is examined as a continuous variable, we see the difference in the distributions, contributing to the median age of controls being 3 years older than the remdesivir patients in our study. For CRP, controls are more skewed towards lower CRP values.

**Supplementary Figure 2: Density plots of baseline continuous measures by treatment group: matched cohort**

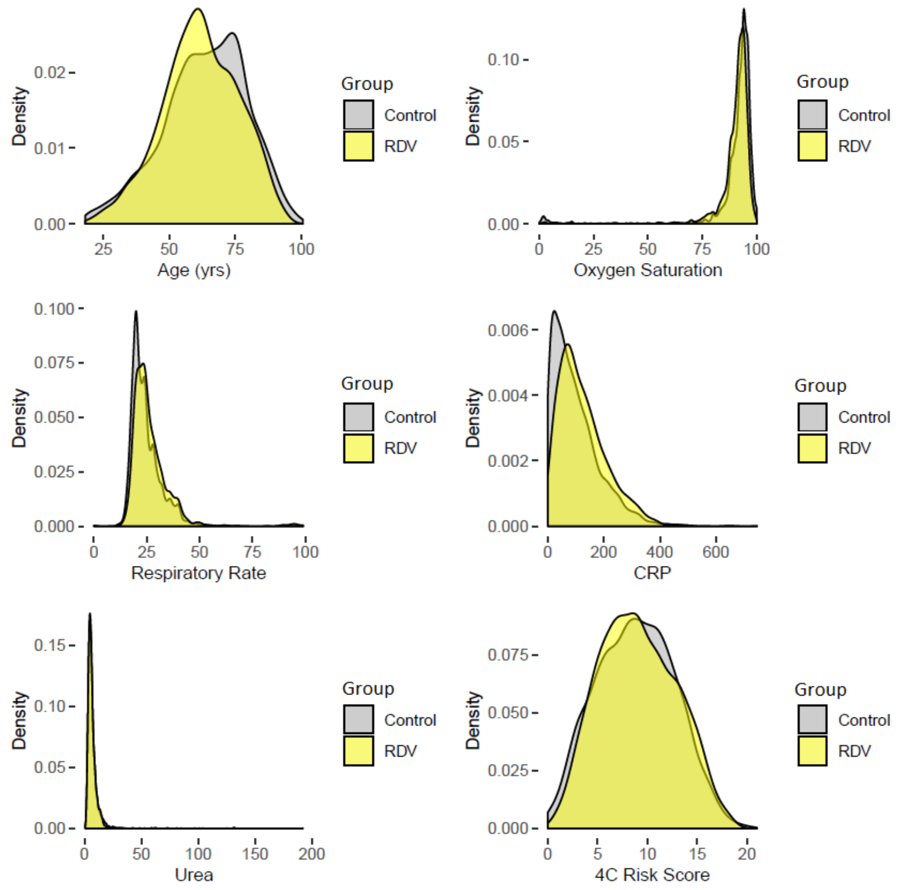

### 9. Treatments received during hospitalisation

Supplementary Table 7 summarises data collected on treatment for each treatment group.

**Supplementary Table 7: Medication received during hospitalisation or at discharge (Matched Cohort)**

| | Remdesivir (n=1,549) | Control (n=4,964) |
|---|---|---|
| **Antiviral agent?** | | |
| Yes | 1,549 (100%) | 150 (3.0%) |
| No | 0 | 4,422 (89.1%) |
| Not known | 0 | 392 (7.9%) |
| **Type of antiviral agent** | | |
| Remdesivir | 1,549 (100%) | 0 |
| IL6 inhibitor | 16 (1.0%) | 47 (0.9%) |
| Lopinavir/Ritonavir | 6 (0.4%) | 17 (0.3%) |
| Chloroquine / Hydroxychloroquine | 1 (0.1%) | 15 (0.3%) |
| Oseltamivir (Tamiflu®) | 0 | 5 (0.1%) |
| Neuraminidase inhibitors | 0 | 4 (0.1%) |
| Other antiviral | 13 (0.8%) | 57 (1.1%) |
| **Remdesivir (days received):** | | |
| 1 | 73 (5.0%) | - |
| 2 | 98 (6.3%) | - |
| 3 | 105 (6.8%) | - |
| 4 | 174 (11.2%) | - |
| 5 | 800 (51.6%) | - |
| >5 | 216 (13.9%) | - |
| Unknown | 83 | |
| **IL6 inhibitor (type)** | | |
| Tocilizumab | 15 (1.0%) | 34 (0.7%) |
| Other | 1 (0.1%) | 13 (0.3%) |
| **IL6 inhibitor (days received):** | | |
| N, Median (IQR) | 15, 2 (1, 2) | 44, 1 (1,2) |
| **Antibiotic** | | |
| Yes | 1,389 (89.7%) | 3,960 (79.8%) |
| No | 124 (8.0%) | 698 (14.1%) |
| Not known | 36 (2.3%) | 306 (6.2%) |
| **Any dexamethasone given?** | | |
| Yes | 1,455 (93.9%) | 3,063 (61.7%) |
| Yes*, at 6mg per day* | 1,375 (88.8%) | 2,874 (57.9%) |
| Yes, but different dose (od) | 64 (4.1%) | 137 (2.8%) |
| Yes: 6mg, but different frequency | 5 (0.3%) | 13 (0.3%) |
| Yes: different dose and frequency | 11 (0.7%) | 39 (0.8%) |
| No | 49 (3.2%) | 1,431 (28.8%) |
| Not known | 45 (2.9%) | 470 (9.5%) |
| **Corticosteroid other than dexamethasone** | | |
| Yes | 145 (9.4%) | 529 (10.7%) |
| No | 1,294 (83.5%) | 3,900 (78.6%) |
| Not known | 110 (7.1%) | 535 (10.8%) |
| **Antifungal agent** | | |
| Yes | 70 (4.5%) | 160 (3.2%) |
| No | 1,414 (91.3%) | 4,397 (88.6%) |
| Not known | 65 (4.2%) | 407 (8.2%) |
| **Off-label / compassionate use medications** | | |
| Yes | 54 (3.5%) | 92 (1.9%) |
| No | 1,400 (90.4%) | 4,270 (86.0%) |
| Not known | 95 (6.1%) | 602 (12.1%) |
| **Interleukin inhibitors** | | |
| Yes | 23 (1.5%) | 21 (0.4%) |
| No | 1,445 (93.3%) | 4,475 (90.1%) |
| Not known | 81 (5.2%) | 468 (9.4%) |
| **Convalescent plasma** | | |
| Yes | 124 (8.0%) | 242 (4.9%) |
| No | 1,346 (86.9%) | 4,266 (85.9%) |
| Not known | 79 (5.1%) | 456 (9.2%) |

### 10. Complications reported in the Outcome CRF

Supplementary Table 8 summarises complications reported for each treatment group.

**Supplementary Table 8: Complications reported at any time during hospitalisation in descending order of overall incidence (Matched Cohort, where complications CRF was completed)**

| | Remdesivir (n=1,504) | Control (n=4,686) | Total (n=6,190) |
|---|---|---|---|
| Viral pneumonia | 1,131 (75.2%) | 2,701 (57.6%) | 3,832 (61.9%) |
| Bacterial pneumonia | 213 (14.2%) | 515 (11.0%) | 728 (11.8%) |
| Hyperglycaemia | 268 (17.8%) | 411 (8.8%) | 679 (11.0%) |
| Acute renal injury/acute renal failure | 127 (8.4%) | 423 (9.0%) | 550 (8.9%) |
| Anaemia | 128 (8.5%) | 375 (8.0%) | 503 (8.1%) |
| Acute respiratory distress syndrome | 172 (11.4%) | 276 (5.9%) | 448 (7.2%) |
| Liver dysfunction | 124 (8.2%) | 242 (5.2%) | 366 (5.9%) |
| Cardiac arrhythmia | 95 (6.3%) | 250 (5.3%) | 345 (5.6%) |
| Pleural effusion | 46 (3.1%) | 184 (3.9%) | 230 (3.7%) |
| Coagulation disorder / disseminated intravascular coagulation | 80 (5.3%) | 145 (3.1%) | 225 (3.6%) |
| Bacteraemia | 38 (2.5%) | 145 (3.1%) | 183 (3.0%) |
| Pulmonary thromboembolism | 45 (3.0%) | 126 (2.7%) | 171 (2.8%) |
| Congestive heart failure | 32 (2.1%) | 98 (2.1%) | 130 (2.1%) |
| Other neurological complication | 20 (1.3%) | 99 (2.1%) | 119 (1.9%) |
| Cardiac arrest | 23 (1.5%) | 89 (1.9%) | 112 (1.8%) |
| Hypoglycaemia | 26 (1.7%) | 67 (1.4%) | 93 (1.5%) |
| Pneumothorax | 20 (1.3%) | 38 (0.8%) | 58 (0.9%) |
| Gastrointestinal haemorrhage | 7 (0.5%) | 51 (1.1%) | 58 (0.9%) |
| Cardiac ischemia | 13 (0.9%) | 45 (1.0%) | 58 (0.9%) |
| Stroke / Cerebrovascular accident | 7 (0.5%) | 45 (1.0%) | 52 (0.8%) |
| Seizure | 5 (0.3%) | 37 (0.8%) | 42 (0.7%) |
| Deep vein thrombosis | 4 (0.3%) | 20 (0.4%) | 24 (0.4%) |
| Cardiomyopathy | 3 (0.2%) | 18 (0.4%) | 21 (0.3%) |
| Cryptogenic organizing pneumonia (COP) | 6 (0.4%) | 13 (0.3%) | 19 (0.3%) |
| Rhabdomyolsis | 3 (0.2%) | 12 (0.3%) | 15 (0.2%) |
| Myocarditis / Pericarditis | 3 (0.2%) | 12 (0.3%) | 15 (0.2%) |
| Bronchiolitis | 3 (0.2%) | 10 (0.2%) | 13 (0.2%) |
| Endocarditis | 3 (0.2%) | 9 (0.2%) | 12 (0.2%) |
| Pancreatitis | 1 (0.1%) | 13 (0.3%) | 14 (0.2%) |
| Meningitis / Encephalitis | 0 | 9 (0.2%) | 9 (0.1%) |

**11. Rationale for chosen approach of outcome analysis methods**

The matching algorithm used in this study created clusters of control patients for each remdesivir patient, and within each cluster, propensity scores were by design similar. There has been debate in the literature as to whether this clustering should be considered in outcome analyses, and further research is needed before a consensus on this matter is reached. In this study, we followed the advice of Stuart [9], who gives two reasons why it is not necessary to account for clustering (or equivalently matched-pairs in 1:1 matching): "*First, conditioning on the variables that were used in the matching process (such as through a regression model) is sufficient. Second, propensity score matching in fact does not guarantee that the individual pairs will be well-matched on the full set of covariates, only that groups of individuals with similar propensity scores will have similar covariate distributions.*" This approach also has the advantage of being simpler to implement, and easier to interpret.

We decided to adjust for two variables that were not defined *a priori*. Viral pneumonia, and use of dexamethasone. An indicator of viral pneumonia during hospitalisation in the complications CRF suggested that the remdesivir group may have been a more ill cohort, despite inclusion criteria and matching on baseline characteristics suggesting that the cohorts were balanced, with similar characteristics and similar disease severity. As stated in our discussion, higher reported viral pneumonia in the remdesivir group is difficult to interpret, and arguably, all our cohort would have had viral pneumonia at baseline, by definition of the inclusion criteria. The CRF was designed as a general tool, and not specifically for COVID-19. We added it as a potential confounder in case this could act as a proxy for an unmeasured baseline factor. Dexamethasone was recorded as given to most patients that were given remdesivir, but not in all controls. Adjusting for this in analyses was designed to enable an estimate of the effectiveness of remdesivir over and above the effect of dexamethasone. This approach is limited, as it does not take into account (a) when dexamethasone was started, and (b) for how long it was given. Clinical opinion among the authors of this paper was that it's prescribing would have been independent of whether patients were given remdesivir, and as an indicator variable it may act as a proxy for identifying patients that were more ill at baseline. Results of models excluding these variables are given below as sensitivity analyses.

**12. Sensitivity analysis of the Primary Outcome**

The following sensitivity analyses were carried out: (1) to check whether adding propensity score as an additional covariate modified effect size or inference – inference was not changed; (2) replacing age, sex and number of comorbidities with propensity score (since this is a function of these as well as other factors), again conclusions did not change rather the adjusted odds ratio was revised down to 0.77, 95%CI: 0.59-1.01, p=0.062; (3) adding a treatment-by-age interaction to this model - this gave a signal as to why the confidence interval is fairly wide – an interaction was found to be significant for the oldest patients (80 plus), moving the point estimate of the odds-ratio in this group close to 1; (4) removing viral pneumonia and/or dexamethasone – inference was not changed.

The unadjusted odds-ratio 95% confidence interval was 0.60-0.88. This is much tighter. All multivariable models resulted in wider confidence intervals than this – and it is likely due to heterogeneity of 14-day mortality among the subgroups represented by fitted covariates. Larger sample sizes are therefore required when adjusting for covariates.

### 13. Clinical Status at day 15

The study inclusion/exclusion criteria were designed such that all patients analysed were at the same clinical status at baseline: hospitalised, requiring supplemental oxygen, but not ventilated. Using the same scoring system as defined in the ACTT-1 trial, this is equivalent to a clinical status score of 5. Therefore, a score of less than 5 at 15 days indicates improvement, and a score of 6 or higher implies worsening. The statistical analysis plan assumed some data could be gathered from daily CRFs to identify whether patients were still on oxygen or required ventilations at 15 days. However, for our cohort, daily CRFs were not generally completed after day 9. The maximum day was day 14, but just 4 controls and 3 remdesivir patients had CRFs completed for this day or the day before. Other data regarding this outcome are collected in the Outcome CRF, though not all details are always complete to enable a clear categorisation. Where a patient could be in more than one category, an average clinical status score was assigned. Clinical statuses 4-6 could not be differentiated at day 15, and there were very small numbers of patients classified as at status 3 or 7 (Supplementary Table 9). We therefore derived the following 5 clinical statuses: (I) Not hospitalised, with no limitations on activities; (II) Not hospitalised, but possibly needing oxygen and activities may be limited; (III) Not hospitalised, but with some limitation on activities and/or requiring home oxygen, OR hospitalised but no longer requiring COVID related medical care; (IV) Hospitalised, requiring ongoing COVID related medical care, but no mechanical ventilation/ECMO; (V) Hospitalised, on mechanical ventilation or ECMO, OR discharged to palliative care, OR died.

**Supplementary Table 9: Number and percentage of patients in each clinical status at day 15, by treatment group**

| | **Remdesivir (n=1,549)** | **Control (n=4,964)** |
|---|---|---|
| Ordinal score at day 15 (+/- 2 days) | | |
| 1: Not hospitalised, and no limitations on activities | 829 (53.5%) | 2,481 (50.0%) |
| 1.5: Not hospitalised, but home oxygen required / limitation on activities unknown. | 182 (11.7%) | 497 (10.0%) |
| 2: Not hospitalised, limitation on activities and/or requiring home oxygen | 95 (6.1%) | 324 (6.5%) |
| 3: Hospitalised, not requiring supplemental oxygen – no longer requiring ongoing COVID related medical care | 0.00% | 1 (0.0%) |
| 5: Hospitalised, requiring ongoing COVID related medical care | 228 (14.7%) | 759 (15.3%) |
| 7: Hospitalised, on mechanical ventilation or ECMO | 2 (0.1%) | 5 (0.1%) |
| 8: Death / palliative discharge | 161 (10.4%) | 637 (12.8%) |
| Unable to classify | | |
| Transferred prior to Day 18 | 17 (1.1%) | 75 (1.5%) |
| Paliative discharge prior to Day 18 | 2 (0.1%) | 20 (0.4%) |
| Insufficient data | 33 (2.1%) | 165 (3.3%) |

Supplementary Figure 3 presents the log-odds of a higher (worse) clinical status compared with a lower one for different dichotomies of classifications (using the classifications as defined above). As the treatment specific lines plotted are an equal distance apart for each dichotomy, this gives strength to the proportional odds assumption in fitting an ordinal logistic regression.

**Supplementary Figure 3: Log-odds of clinical status by treatment group for different dichotomy comparisons**

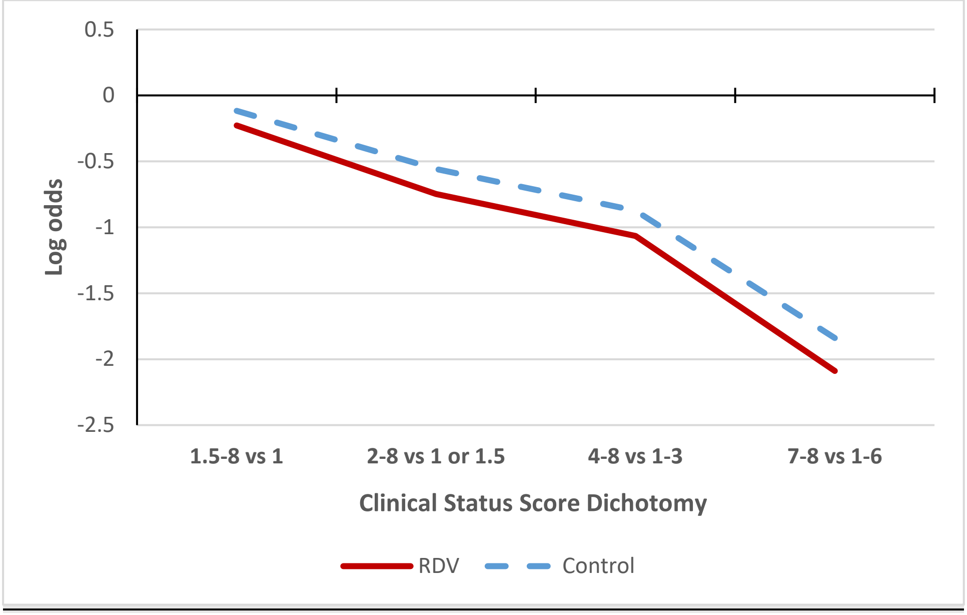

### 14. Checks on data quality regarding ventilation status

Checks were made to assess whether observed differences could possibly be due to a disparity between quality of data collection between the two groups. In 3.4% of remdesivir patients, NIV use was unknown, compared with 8.1% of controls – but even if these unknowns all received NIV, we would still see a large difference in NIV rates. Where the outcome was known, this was recorded in the Outcome CRF in 98% of cases, and regardless of treatment group. By design in the protocol, remdesivir patients have more daily CRFs completed, but daily CRFs identified instances of ventilation mode missing from the Outcome CRF in only 2% of cases – irrespective of group. We are unable to find baseline differences that might explain the higher rates of NIV in the remdesivir group.

### 15. Discussion of Secondary Outcomes

28-day mortality was significantly associated with remdesivir, although the confidence interval for the adjusted odds-ratio is wide, indicating a substantial level of uncertainty about the true effect size.

Rate of recovery was found to be time dependent. It is of interest that the control group tended to recover faster, but that those who did not recover within one week of baseline then had a slower recovery time compared with remdesivir patients. The remdesivir group had slower initial recovery –

this could be explained if these patients had a delayed discharge during the treatment phase, or if the treatment has a latent effect. It may also indicate that there is an unmeasured factor associated with the control group explaining faster recovery. Clinicians may have given priority for remdesivir use to patients that they expected would make a slower recovery, or for whom some risk or poor outcome was identified that we could not capture, yet the predictive 4C Mortality Scores at baseline were similar between groups.

Clinical status at day 15 could not be measured with the level of precision that had been pre-planned. Instead of an 8-point ordinal scale, we were limited to deriving a 5-point scale which was similar. However, this means that our analysis is not directly comparable with the ACTT-1 trial results, which detected a significant benefit in remdesivir over placebo in clinical status at day 15. In our study, we find no significant association of remdesivir with clinical status at this time-point – this is no surprise, as we have observed that in our cohort, Remdesivir appear to be associated more with benefit at 28 days.

Use of ventilation was more prevalent in the remdesivir group – especially non-invasive ventilation, in which rates were almost three times higher compared with controls. There is no scope in this study to investigate why this was the case. We also do not know when ventilation was initiated, nor the duration of non-invasive forms. There may be an unmeasured bias in the determinant of who received remdesivir, and if this was also associated with likelihood of ventilation, this would explain the observed differences. It may be that if there is a latent effectiveness for remdesivir, some ventilation may be required in the short-term. Existing clinical trial evidence does not indicate that it is likely that remdesivir use would have caused higher rates of ventilation.
